# Within-host SARS-CoV-2 viral kinetics informed by complex life course exposures reveals different intrinsic properties of Omicron and Delta variants

**DOI:** 10.1101/2023.05.17.23290105

**Authors:** Timothy W. Russell, Hermaleigh Townsley, Sam Abbott, Joel Hellewell, Edward J Carr, Lloyd Chapman, Rachael Pung, Billy J. Quilty, David Hodgson, Ashley S Fowler, Lorin Adams, Christopher Bailey, Harriet V Mears, Ruth Harvey, Bobbi Clayton, Nicola O’Reilly, Yenting Ngai, Jerome Nicod, Steve Gamblin, Bryan Williams, Sonia Gandhi, Charles Swanton, Rupert Beale, David LV Bauer, Emma C Wall, Adam Kucharski

## Abstract

The emergence of successive SARS-CoV-2 variants of concern (VOC) during 2020-22, each exhibiting increased epidemic growth relative to earlier circulating variants, has created a need to understand the drivers of such growth. However, both pathogen biology and changing host characteristics – such as varying levels of immunity – can combine to influence replication and transmission of SARS-CoV-2 within and between hosts. Disentangling the role of variant and host in individual-level viral shedding of VOCs is essential to inform COVID-19 planning and response, and interpret past epidemic trends. Using data from a prospective observational cohort study of healthy adult volunteers undergoing weekly occupational health PCR screening, we developed a Bayesian hierarchical model to reconstruct individual-level viral kinetics and estimate how different factors shaped viral dynamics, measured by PCR cycle threshold (Ct) values over time. Jointly accounting for both inter-individual variation in Ct values and complex host characteristics – such as vaccination status, exposure history and age – we found that age and number of prior exposures had a strong influence on peak viral replication. Older individuals and those who had at least five prior antigen exposures to vaccination and/or infection typically had much lower levels of shedding. Moreover, we found evidence of a correlation between the speed of early shedding and duration of incubation period when comparing different VOCs and age groups. Our findings illustrate the value of linking information on participant characteristics, symptom profile and infecting variant with prospective PCR sampling, and the importance of accounting for increasingly complex population exposure landscapes when analysing the viral kinetics of VOCs.

## Background

Successive SARS-CoV-2 variants of concern (VOC) have exhibited step-wise increases in relative epidemic growth rates (1–3). The selective advantage of successive VOCs are the result of complex epistatic relationships arising from combinations of mutations occurring in the viral genome. Selective advantage is frequently associated with mutations in the region encoding the spike protein. To increase the growth rate of a VOC within a community, these mutations must confer improved evasion of antibody-mediated immunity, increased intrinsic transmissibility or a combination of both. Understanding how inter-related factors including pathogen biology and host characteristics – such as pre-existing immunity – impact on replication and transmission of SARS-CoV-2 is essential to inform ongoing COVID-19 planning and response.

Over the course of a SARS-CoV-2 infection, the viral load in the nasopharynx increases after infection until reaching a peak and then declining. The dynamics of the viral load over time are known as viral kinetics. Viral kinetics are quantified by performing RT-PCR on nasal swabs, this returns a cycle threshold (Ct) value that measures the concentration of viral RNA in the nasopharynx (4). Thus, the cycle thresholds inversely correlate with higher RNA viral concentrations. Such values measure viral RNA in the nasopharynx, which correlate well with live virus as measured by PFU/ml in the early stages of infection (4). Viral kinetics will vary between individuals due to host factors such as age, sex, or prior immunity, or due to changes in the pathogen related to the emergence of new VOCs (4,9–14).

Viral kinetics may change between variants and epidemic waves (5,15), necessitating updating of recommendations as new variants become dominant. However, as population immunity accumulates via infection and vaccination, it will become harder to generalise descriptive insights from one population to another. Studies that can identify factors influencing viral kinetics and likely infectiousness are therefore crucial for developing up-to-date public guidance and understanding risk of transmission for patients.

Changes in viral kinetics have implications for transmission since higher viral loads are frequently associated with increased host transmissibility. There are several reasons higher transmissibility could be observed after the emergence of a new VOC, including: higher immune escape; faster initial replication; higher peak viral load; longer period of viral shedding; or a different symptom profile, and hence different host behaviour. At this point in the COVID-19 pandemic everyone has a unique life course exposure history due to their past infections and vaccinations. These complex life course exposure histories make it challenging to determine whether changes in observed viral kinetics are due to host factors such as prior immunity or changes in the pathogen itself. Studies that can separate such factors influencing viral kinetics, and hence likely infectiousness, are therefore crucial for developing up-to-date public guidance and understanding risk of transmission for patients.

Previous studies have sought to relate within-host viral kinetics with other measurable properties such as disease severity, mortality, VOC, and the probability of onward transmission (9, 10, 16, 4, 12–14). However, they have often had one of two key limitations. Either they have used serial sampling following an initial diagnostic swab, conducted only after symptom onset, thus missing the early rates of viral replication (17). Or, they used single sampling with Ct values that correspond to a single time point per individual, meaning that variation in the viral load over the course of an infection is not distinguishable from the variation in viral load between individuals (2, 21). As a result, any conclusions about differences in Ct value between individuals may be an artefact of factors such as diagnostic protocols or underlying epidemic dynamics.

Repeatedly testing many individuals throughout their infection, including in the early pre-symptomatic period, makes it possible to separate individual Ct dynamics over time from the between-individual variation in viral load (5, 19). In cohort studies where individuals have been repeatedly tested to infer the viral dynamics of their infection, this between-individual variation has previously been compared to single characteristics, including: the infecting VOC (5,6,9), age (9, 11), and vaccination status (8, 13). However, there can be considerable confounding between different characteristics. For example, each sequential VOC has spread against a different background of overall population vaccine coverage, including numbers of doses received, confounding any comparisons based only on VOC. Failure to adjust for such confounding factors risks attributing differences in viral load to different VOCs that are in fact associated with factors that influence viral replication, such as the presence of vaccine-induced immune effectors such as neutralising antibody levels. Accounting for both inter-individual variation in Ct values and complex host characteristics – such as vaccination status, exposure history and age – is therefore crucial for obtaining reliable estimates of the relationship between variants of concern (VOCs) and viral kinetics (21).

To address this complexity, we developed a Bayesian hierarchical modelling framework that estimates the unobserved Ct trajectories for each individual, using data from a subset of a prospective cohort undergoing weekly occupational health PCR screening for SARS-CoV-2. As well as accounting for different covariates jointly, this modelling approach pools data across individuals to identify the main drivers of variation in Ct dynamics and estimate the accompanying uncertainty. Using this model, we examined the relationship between Ct dynamics and different VOCs, including timing and peak viral loads, and quantified the impact of key individual-level characteristics on these dynamics.

## Methods

### Clinical cohort composition

The Legacy study is a prospective observational cohort study of healthy adult volunteers undergoing weekly occupational health PCR screening for SARS-CoV-2 at University College London Hospitals or at the Francis Crick Institute (NCT04750356) (22,23). The Legacy study was approved by London Camden and Kings Cross Health Research Authority Research and Ethics committee (IRAS number 286469) and is sponsored by University College London Hospitals. Participants provided baseline demographics including age, sex, and medical comorbidities, with data collection on subsequent vaccine doses and COVID-19 infections. Participants reporting either a positive test result for SARS-CoV-2, or onset of symptoms consistent with COVID-19, were sent same-day swabs via courier up to day 10 post positive test or symptom onset. They received further swabs every 2-3 days until day 10 (**Figure 1A**). All participants had a face-to-face post-infection study visit before day 30 post-infection, at which point an additional swab was collected, and a symptom diary was completed (24). We linked diagnostic swab data with earlier swab data from the occupational screening pipeline at the Francis Crick Institute, enabling estimation of the pre-symptomatic period between the last reported negative screening swab and onset of SARS-CoV-2 infection.

**Figure 1.**
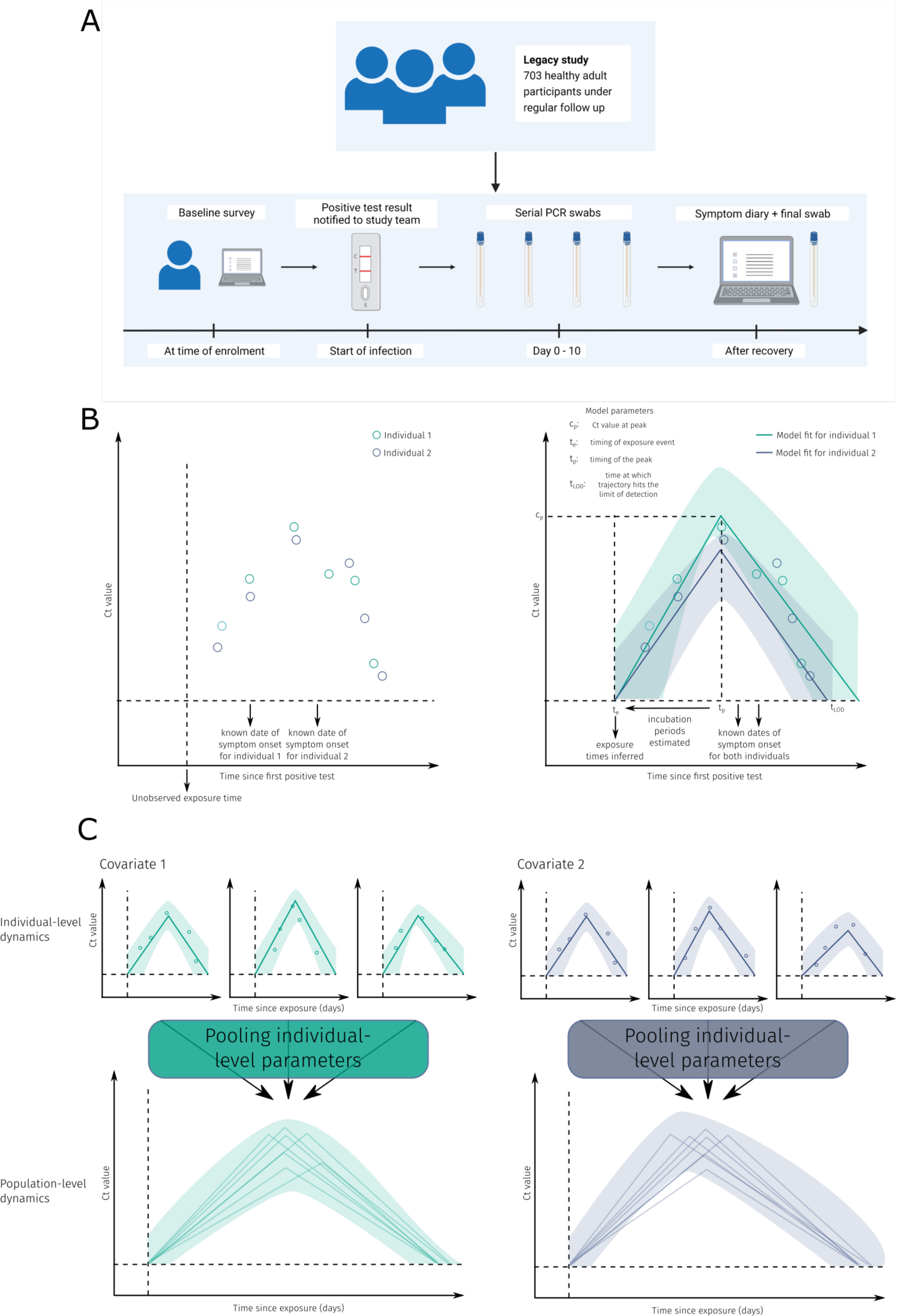
Schematic of the study design and modelling procedure. **A.** Schematic of the cohort and data collection procedure used for the subset of the Legacy study. **B.** Typical Ct value data and model fits for two representative individuals with different covariates. **C.** Schematic of the individual-level and covariate-level model fits. The model fits each participants viral kinetics and pools the estimates in a statistically robust manner.

We included infection episodes reported by Legacy study participants which were associated with at least two available positive RT-PCR results. The total size of the cohort up to the maximum date considered was 680 participants, of which 135 had two positive results and as such, were included in this analysis. For each infection episode, whole genome sequencing or a combination of date of infection and viral genotype was used to assign the VOC that caused the infection. Infection episodes occurred between 3^rd^ June 2021 and 25^th^ April 2022.

### SARS CoV-2 RT-PCR and viral sequencing

Viral RNA was extracted from nasopharyngeal swabs and analysed by RT-PCR using methods previously described in Aitken et al. (25). ORF1ab, N and S gene Ct values were used for this analysis, with an in-model adjustment for gene type. We found strong correlation between the Ct values at the different gene targets (**Figure S1**). Therefore, including all Ct values available increased statistical power for each individual. Viral RNA from positive swabs was prepared for whole-genome sequencing using the ARTIC method (https://www.protocols.io/view/ncov-2019-sequencing-protocol-v3-locost-bh42j8ye) and sequenced on the ONT GridION platform to >30k reads / sample. The data were demultiplexed and processed using the viralrecon pipeline (https://github.com/nf-core/viralrecon) (25).

### Estimating the incubation period of SARS-CoV-2 variants

In total, 126/140 (91.4%) of infection episodes had symptoms reported, reported by their date of symptom onset (**Table 1**). Participants were a subset of the wider post-vaccine infection cohort within the Crick, where serial sampling data were available (24). We used the difference between the estimated time of exposure and the time of symptom onset for each individual to fit two parameters of an assumed log-normal form of the incubation period for SARS-CoV-2 (**Figure 1B-C**). We specified semi-informative priors for both parameters using existing estimates of the incubation period (26).

**Table 1.** Number of participants in each covariate category used in our study. Covariate categories used in the baseline model include: VOC status, symptom status, total number of exposures at time of infection and age group. The other covariates are investigated in one of the various alternative models considered.

| Table 1: Demographic details of participants<sup>2</sup> |  |  |  |
| --- | --- | --- | --- |
| Characteristic | Delta, N = 34<sup>1</sup> | Omicron BA.1, N = 48<sup>1</sup> | Omicron BA.2, N = 58<sup>1</sup> |
| Female | 18 (53%) | 33 (68%) | 39 (67%) |
| Male | 16 (47%) | 15 (32%) | 19 (33%) |
| Median age (years) [IQR] | 35 [22—48] | 41 [21—61] | 34 [18—50] |
| Number of doses at time of infection [* infection commenced within 0-14d of dose] |  |  |  |
| 2* | 2 (3.7%) | 0 (0%) | 0 (0%) |
| 2 | 32 (59.3%) | 16 (30.1%) | 1 (1.7%) |
| 3* | 18 (33.3%) | 5 (9.4%) | 0 (0%) |
| 3 | 2 (3.7%) | 32 (60.4%) | 57 (98.3%) |
| Dose 1 |  |  |  |
| AZD1222 | 16 (47.1%) | 14 (29.2%) | 17 (29.3%) |
| BNT162b2 | 18 (52.9%) | 32 (66.7%) | 36 (62.1%) |
| mRNA1272 | 0 (0%) | 2 (4.2%) | 4 (6.9%) |
| others | 0 (0%) | 0 (0%) | 1 (1.7%) |
| Dose 2 |  |  |  |
| AZD1222 | 16 (47.1%) | 14 (29.2%) | 16 (27.6%) |
| BNT162b2 | 18 (52.9%) | 32 (66.7%) | 37 (63.8%) |
| mRNA1272 | 0 (0%) | 2 (4.2%) | 4 (6.9%) |
| others | 0 (0%) | 0 (0%) | 1 (1.7%) |
| Dose 3 |  |  |  |
| BNT162b2 | 10 (29.4%) | 34 (66.7%) | 56 (96.6%) |
| mRNA1272 | 2 (5.9%) | 16 (31.4%) | 2 (3.4%) |
| N/A | 22 (64.7%) | 0 (0%) | 0 (0%) |
| Unknown | 0 (0%) | 1 (0.02%) | 0 (0%) |
| Total number of exposures at time of included infection episode |  |  |  |
| Characteristic | Delta, N = 34<sup>1</sup> | Omicron BA.1, N = 48<sup>1</sup> | Omicron BA.2, N = 58<sup>1</sup> |
| 3 | 20 (58.9%) | 11 (22.9%) | 1 (1.7%) |
| 4 | 11 (32.4%) | 29 (60.4%) | 41 (70.7%) |
| 5+ | 3 (8.9%) | 8 (16.7%) | 16 (27.6%) |
| Joined study before infection episode? |  |  |  |
| No | 14 (42%) | 33 (52%) | 22 (38%) |
| Yes | 19 (58%) | 30 (48%) | 36 (62%) |
| Median days since dose prior to infection [IQR] | 121 [83.5—184] | 76 [34.0—147] | 93 [69.5—113] |
| Self-reported symptom severity |  |  |  |
| grade I | 12 (38%) | 27 (44%) | 19 (35%) |
| grade II | 11 (34%) | 19 (31%) | 28 (51%) |
| grade III | 0 (0%) | 2 (3.2%) | 0 (0%) |
| asymptomatic | 9 (28%) | 14 (23%) | 8 (15%) |
| Unknown | 2 | 1 | 3 |
| Self-reported duration of symptoms [IQR] | 10 [7-14] days | 10 [8-14] days | 11 [8-15] days |
| Virus sequenced | 32 (94%) | 56 (89%) | 32 (55%) |

<sup>1</sup>n (%); Median [25%-75%]

### Estimating viral kinetics by variant and host characteristics

We constructed a Bayesian model that jointly infers individual-level Ct trajectories, population (covariate)-level trajectory parameters, parameters of a log-normally distributed incubation period and multiplicative effect size parameters following a linear regression. We regress against several covariates for both the population-level Ct trajectory and incubation period parameters. Covariates used are described in detail in the next section. The model allows for the addition (or removal) of covariates through a formula interface.

For the individual-level Ct trajectories, we extended and modified an existing model of individual-level cycle threshold dynamics over the course of entire infections of any respiratory disease measurable by RT-PCR test and applied this to our cohort of participants infected with SARS-CoV-2 (5,6). The individual-level dynamics are modelled with a piecewise linear curve, on a logarithmic scale, with a single breakpoint, where the breakpoint represents the peak of the trajectory (**Figure 1B**). The logarithmic scale corresponds to assuming an exponential increase in viral load from the point of exposure until the peak, then an exponential decrease at a different rate to the point of clearance.

Each individual trajectory was parameterised using four parameters with the following epidemiological interpretations, each corresponding to events that were not directly observed: the timing of initial exposure event, timing of the peak (lowest) Ct value, Ct value at the peak and the timing the trajectory hits the limit of detection (LOD, highest Ct value) (**Figure 1B-C**), which for this assay was Ct = 40. The reported LOD is a censored value, meaning in theory a more sensitive machine could detect SARS-CoV-2 at higher Ct values. As such, we also estimate a true (censored) LOD for each individual, both at the start and end of each infection episode. We report differences in the peak Ct value, the timing of the peak and the timing trajectories hit the censored LOD.

The mechanistic model generated the expected Ct value at a given time since exposure for each individual, which was compared to the measured Ct values using a normally distributed observation process. The variance parameter of this term was shared across all individuals and represents the measurement error from the RT-PCR sampling and testing process. Given that we fit to data at three gene targets (ORF1ab, S and N-gene targets), we include a term in the Bayesian linear regression adjusting for differences in gene target (**Figure S1**).

Given the complex nature of the exposure history of each individual (**Figure 2A**), we adjusted for the time since the last exposure for each individual. We did so by calculating the time since the last infection or vaccination (whichever was sooner) for each individual as well as estimating effect sizes for the total numbers of exposures for each individual. This adjustment was performed in the Bayesian multiple linear regression component of the model and as such it did not include explicit antibody dynamics.

**Figure 2.**
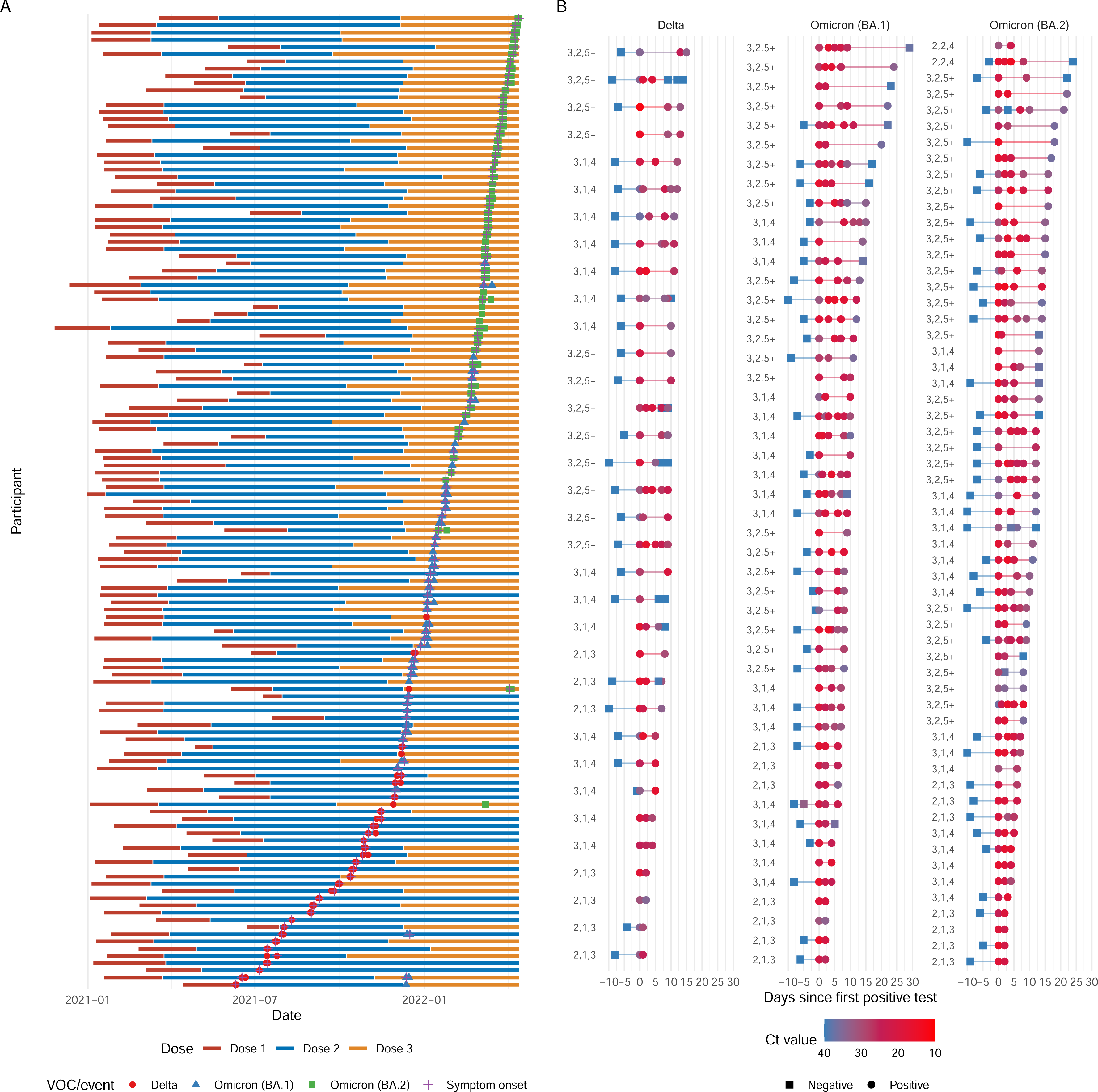
Summary of longitudinal cohort dataset analysed. **A.** Ct value distribution for all participants, stratified by the interaction between the number of vaccines and the infecting VOC, the interaction between the number of infections and the infecting VOC and finally by the four covariate categories used throughout the study: VOC, symptom status, number of exposures and age. **B.** The individual-level Ct data, including the timing of each test, the Ct value of the result, given by the colour and the number of vaccinations, infections, and the total exposures for each participant on the left of their individual timeline.

### Covariate categories and regression reference choices

The categories of covariates used were: VOC status (Delta, Omicron BA.1 or Omicron BA.2), symptom status (symptomatic or asymptomatic), age group (20–34, 35–49 and 50+), total number of antigenic exposures (3, 4 or 5+; calculated as the sum of infections and vaccine doses) and time (days) since the last exposure (see Table 1 for full description of the number of individuals in each category).

In the linear regression framework, values for VOC status, symptom status, age group and numbers of exposures needed to be chosen for the reference case against which, the other covariates were regressed. Symptomatic, Omicron BA.1 individuals, aged between 35––49 years with 4 exposures were chosen as the baseline case for both interpretability and statistical power, as this combination produced the largest reference group of individuals.

After fitting the model, we used the covariate-level posterior distributions to simulate 10,000 Ct trajectories for each covariate included in the analysis, and summarised the trajectories by calculating the median, 50% and 95% credible intervals for each covariate. Details and comparisons of alternative models — using either different model structures or differences in the regression formula — can be found in the supplementary material (**Figures S10, S11, S12 & S13**).

### Data and code availability

The dataset containing the Ct values at the three gene targets considered for the 140 infection episodes, symptom onset times and covariate data, along with the code to re-run the analysis can be found at this publicly available repository: https://github.com/thimotei/legacy_ct_modelling.

## Results

### Differences in peak Ct value by variant and host characteristics

There was considerable variation in both individual characteristics and Ct dynamics across the cohorts (**Figure 2B**). Applying our Bayesian model to these data, we estimated that the Ct trajectories for participants in the reference regression group (Omicron BA.1, symptomatic, 4 exposures and aged 35—49 years) peaked at 16.1 (95% Credible Interval [CrI]: 14.9—17.2) (**Figure 3A, Table S1**). In comparison, we estimated that the trajectories of the Delta and BA.2-infected subsets have peak Ct values of 15.1 (95% CrI: 13.8—16.5) and 15.0 (95% CrI: 13.9—16.1) respectively (**Figure 3A, Table S1**), resulting in differences of 0.93 (95% CrI: −0.440—2.30) and 1.00 (95% CrI: −0.081—2.20) Ct values from baseline individuals (respectively). We estimated that asymptomatic individuals have a peak Ct value of 17.0 (95% CrI: 15.3—18.7) — a peak Ct value 0.94 (95% CrI: −0.90—2.8) higher than baseline individuals (**Figure 3B, Table S2**) — corresponding to a lower peak viral load. We find that individuals with 3 exposures in total have a peak Ct value of 16.7 (95% CrI: 15.1—18.3). These individuals and the baseline individuals have peak Ct values 1.2 (95% CrI: −0.58—3.1) and 1.9 (95% CrI: −0.5—3.2) lower (respectively) than individuals with 5+ exposures (who have a peak Ct value of 17.9 (95% CrI: 16.5—19.4)), corresponding to substantially higher viral load for both 3- and 4-exposure individuals (**Figure 3C, Table S3**). Estimates for the 20—34 age group are almost identical to the estimates for baseline individuals. However, 50+ year olds have peak Ct value estimates of 17.7 (95% CrI: 16.4—19.0), 1.6 (95% CrI: 0.25—3.1) Ct values higher than baseline individuals (**Figure 3D, Table S4**).

**Figure 3.**
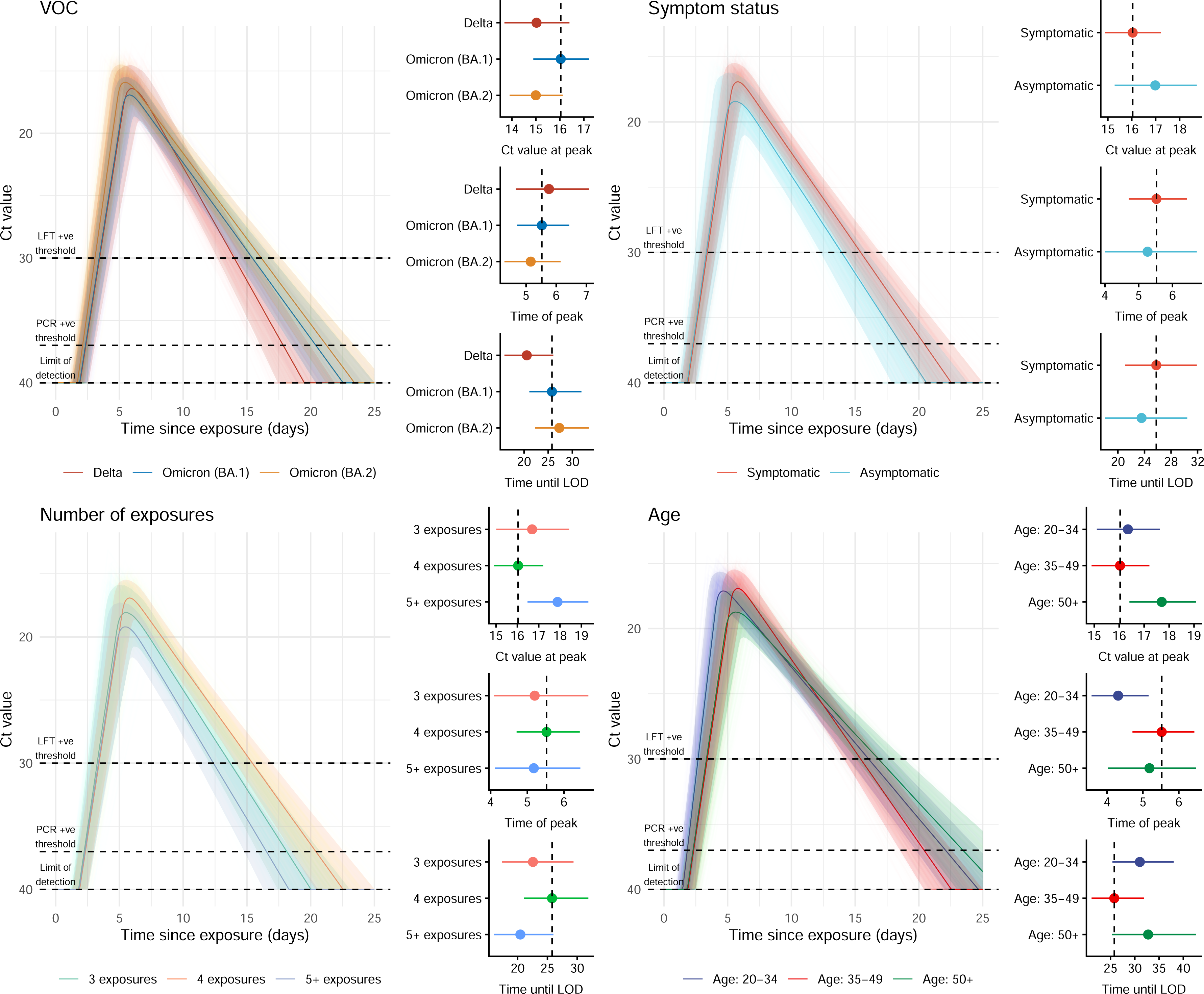
Fitted covariate-level cycle threshold trajectories and the differences in the parameter values stratified for each covariate. **All left panels:** Model fits of the viral kinetics over the course of whole infections, beginning from the inferred exposure time. After fitting the model, we simulated 10,000 trajectories, using parameters sampled from the inferred posterior distributions for each covariate considered. We then calculate the median and 95% credible interval for each covariate and plot the resulting trajectories as coloured regions, with a sample of 100 trajectories plotted transparently beneath. **All right panels:** the median and 95% estimates of the inferred parameter values for the peak Ct value, the timing of the peak and the time at which the trajectory hits the LOD. Dashed line shows the value of the baseline participants inferred parameter values. **A.** Trajectories and parameter values for the VOCs considered. **B.** Trajectories and parameter values for the symptom statuses considered. **C.** Trajectories and parameter values for the numbers of exposures considered. **D.** Trajectories and parameter values for the age groups considered.

### Differences in timing by variant and host characteristics

We estimated that the Ct trajectories in the baseline group (BA.1 infections in 35-49yo after three vaccinations) peaked at 5.6 days after exposure (95% CrI: 4.7—6.5) (**Figure 3A, Table S1**). Delta Ct trajectories peaked at 6.0 days after exposure (95% CrI: 4.9—7.3) and BA.2 trajectories peaked at 5.2 days after exposure (95% CrI: 4.4—6.2), 0.44 (95% CrI: −0.75—1.7) days later and 0.33 (95% CrI: −1.4—0.77) days sooner than the baseline participants respectively (**Figure 3A, Table S1**). Trajectories of asymptomatic participants peaked 5.2 (95% CrI: 4.0—6.7) days after exposure, 0.4 (95% CrI: −1.7—1.1) days sooner than participants in the baseline group (**Figure 3B, Table S2**). We did not find a substantial difference in the timing of the peak for differences in the total numbers of exposures (**Figure 3C, Table S3**). Lastly, we found that Ct trajectories for 20–34 yo and 50+ yo individuals both peak sooner than the baseline group, at 4.2 (95% CrI: 3.5—5.1) and 5.2 (95% CrI: 4.0—6.6), 1.3 (95% CrI: 0.42—2.2) and 0.36 (95% CrI: −0.93—1.5) days sooner than baseline individuals respectively (**Figure 3D, Table S4**).

We then estimated the times at which Ct trajectories reached the thresholds of Ct = 37 or Ct = 30, approximating the limits of detection by PCR or a rapid test that can only detect high Ct values. For PCR positivity (PCR+), baseline trajectories crossed the PCR+ threshold at 2.3 (95% CrI: 1.5—2.9) days after exposure and would have remained PCR+ until 20.3 (95% CrI: 17.3—22.9) days after exposure. We estimated that Delta trajectories would turn PCR+ at an almost identical time to the baseline trajectories, but would only remain PCR+ until 17.9 (95% CrI: 14.8—20.4) days after exposure. We estimated that BA.2 infections would turn PCR+ 2.1 (95% CrI: 1.3—2.8) days after exposure and would remain PCR+ until 21.1 (95% CrI: 18.0— 23.8) days after exposure. Trajectories for all other covariates were estimated to turn PCR+ at very similar times to the baseline trajectories. However, we estimated substantial variation in the duration of PCR positivity for the other covariate categories. Trajectories would become PCR-for participants with 3 or 5+ total antigenic exposures sooner than baseline individuals; specifically at 17.8 (95% CrI: 14.3—20.8) and 16.4 (95% CrI: 13.7—18.8) days after exposure respectively. Lastly, we found that trajectories for participants in age groups 20–34 and 50+ would become PCR-later than reference individuals, at 22.1 (95% CrI: 18.2—25.0) and 23.2 (95% CrI: 18.8—25.0) days after exposure respectively. Estimated rapid test positivity timings are reported in the Supplementary Material.

### Summary of estimated variations in Ct dynamics by variant and host characteristics

Differences within and between all four covariate categories were difficult to characterise precisely, given the complex nature of the exposure history of this dataset. However, some patterns emerged. To summarise, compared to reference individuals, we found: lower peak Ct values for Delta and BA.2 infected-individuals; a later peak timing for Delta infected-individuals and an earlier peak timing for BA.2 infected; Delta infected-individuals reaching the LOD sooner than BA.1 or BA.2 infected-individuals; a substantially higher peak Ct value for individuals with 5+ exposures — compared to reference individuals and individuals with 3 exposures — consistent with lower viral loads and higher levels of immunity for those individuals (22); higher peak Ct values for asymptomatic individuals, consistent with previous estimates and consistent with lower viral loads in asymptomatic individuals throughout their infections (11, 27) and lastly, the timing of the peak Ct value occurred sooner for asymptomatic individuals, consistent with previous estimates (11). However, we estimated that peak Ct values, timing of peak and LOD did not follow a consistent pattern in our dataset across the different age groups. This could be due in part to low statistical power for the highest age group and/or the lack of explicit inclusion of complex underlying antibody dynamics in our model.

### Estimating the relationship between timing of peak shedding and symptom onset

Combining the fitted Ct trajectories with incubation period estimates, we estimated the temporal dynamics of viral shedding and their relation to the timing of symptom onset for each covariate. To compare dynamics, we selected a Ct threshold relating to a time point corresponding to comparatively high viral load (Ct = 20). We found that individuals infected with BA.2 reach the assumed Ct threshold on their upwards trajectory earlier than those infected with BA.1 and that BA.1 individuals reached the Ct threshold earlier than Delta infected individuals. Similarly, using our incubation period estimates, we estimated that symptom onset was earlier for BA.2-infected individuals than for BA.1-infected individuals, and earlier for BA.1-infected individuals than for Delta-infected individuals (**Figure 4**).

**Figure 4.**
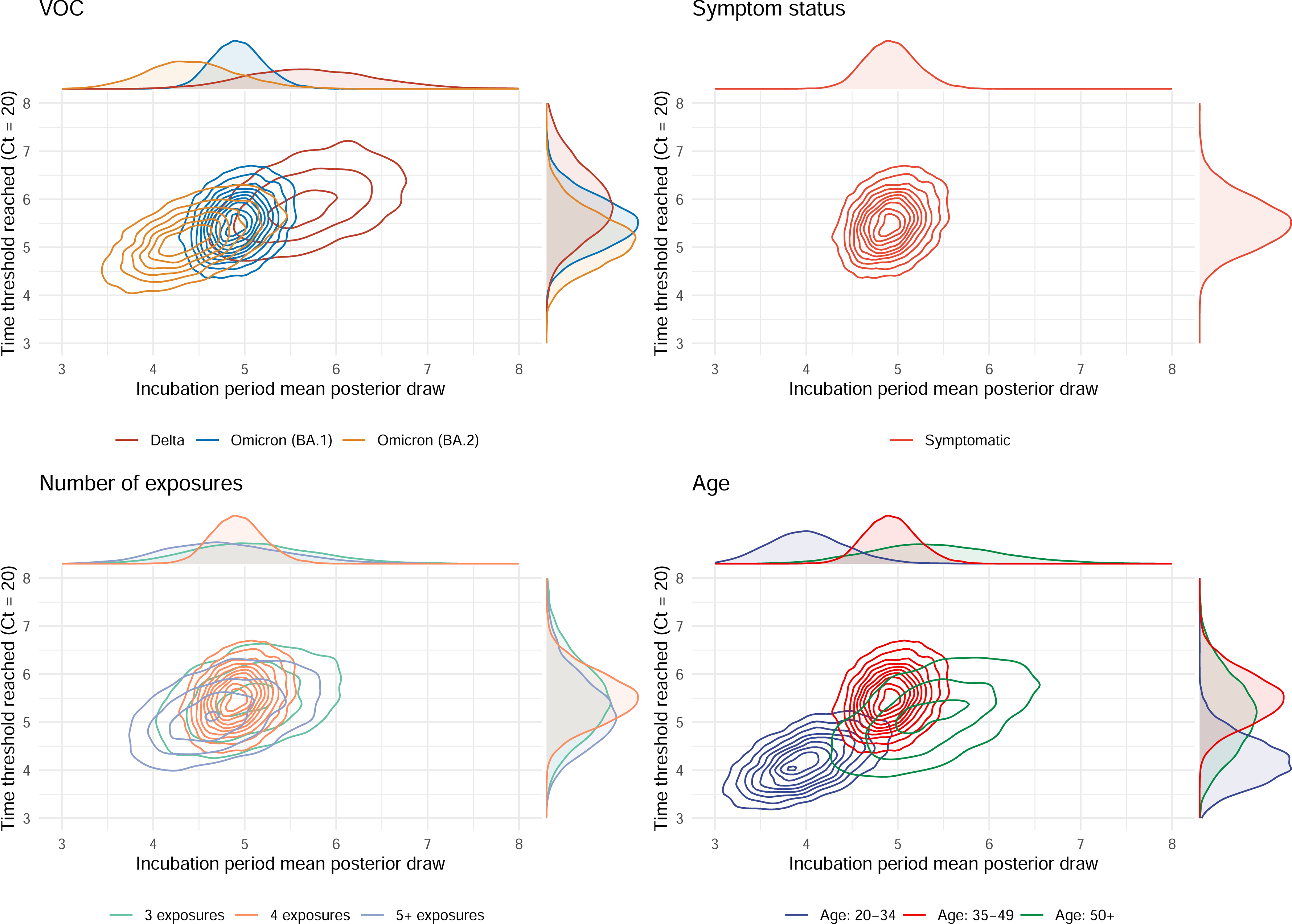
Bivariate density plots of median incubation periods against the time at which trajectories surpassed an assumed Ct value corresponding to high viral loads, for each covariate. **All panels**: We plot the posterior distributions for the estimated incubation periods, stratified by all covariates considered, against the time at which the simulated Ct trajectories, as shown in Figure 3, cross an assumed Ct value threshold for the first time (Ct value = 20). This Ct value represents a timepoint in the covariate-level trajectories of high viral load**. A.** Parameter values for the three VOCs considered. **B.** Parameter values corresponding to symptomatic infections. **C.** Parameter values corresponding to the numbers of exposures considered. **D.** Parameter values for the age groups considered. An alternative Ct value threshold (Ct = 25) was considered in an analogous analysis in the Supplementary Material (**Figure S14**).

Estimates stratified by VOC revealed that these two timescales have become shorter as variants have emerged (**Figure 4A**). For example, the marginal estimates for the time at which the Ct trajectories reached the assumed threshold move from 5.2 (95%: 3.5—6.6) days to 4.9 (95%: 3.6—6.0) days to 4.5 (95%: 3.2—5.6) for the VOCs in the order in which they evolved: Delta → BA.1 → BA.2. Similarly, the incubation periods are 5.7 (95%: 4.0—7.2) days to 4.9 (95%: 4.2—5.5) days to 4.4 (95%: 3.2—5.5) days for the same ordering (**Figure 4A**). As asymptomatic individuals do not have an incubation period, such estimates by symptom status were only possible for symptomatic individuals, which were used as the baseline set of individuals and therefore match those of the BA.1 estimates (**Figure 4B**). We did not find substantial deviations from the reference group when investigating this relationship by total numbers of antigenic exposures (**Figure 4C**). Lastly, when stratified by age group, we estimated the time trajectories pass the assumed threshold moves from 4.9 (95%: 3.1—6.4) days to 4.9 (95%: 3.6—6.0) days to 3.8 (95%: 2.7—4.7) days, and the onset of symptoms occurs at 5.4 (95%: 3.7—6.9) days to 4.9 (95%: 4.2—5.5) to 4.0 (95%: 3.0—4.9) days when age groups are reported by decreasing order: 50+ → 35—49 → 20—34 (**Figure 4D**). Our estimated incubation periods, sampled using both the inferred mean and standard deviation parameters are included in the supplement (**Figure S15).**

## Discussion

We estimated differences in viral shedding dynamics over the course of infections with different SARS-CoV-2 variants of concern by combining data from a large cohort of healthy adults with a statistical model that accounted for both individual- and group-level variation in Ct value. Adjusting for a changing background of host characteristics over time, we identified differences in Ct kinetics that were associated specifically with VOC status, and found that Delta and BA.2 infections tended to peak at higher viral loads compared to BA.1 infections. We also found that age and number of prior exposures had a strong influence on Ct peak, with lower shedding among older individuals and those who had at least five exposures to vaccination and/or infection. Moreover, we found evidence of a correlation between the speed of early shedding and duration of incubation period when comparing different VOCs and age groups.

The unique dataset and model developed here allow us to expand upon previous findings from recent studies that investigated specific aspects of viral kinetics of SARS-CoV-2 infections, both in a human challenge model (4) and in the community (7, 8, 11–13, 15, 31, 32). First, we collected serial swabs from participants in a large, well-defined cohort, which is broadly representative of the UK population, and who were undergoing regular surveillance RT-PCR testing. Second, we captured both asymptomatic and symptomatic infections with high confidence, linked to the last negative test, in comparison with other studies (27) that relied upon self-initiated symptom-based testing. This knowledge of prior negative test timings allowed us to estimate the duration of pre-symptomatic infection with greater confidence, and directly compare the peak and trajectory of symptomatic and asymptomatic infections. One of the major limitations of most studies of SARS-CoV-2 infection has been the unknown time between infection and symptom onset. While the human challenge model approach has provided detailed information about viral kinetics following inoculation, participants were unvaccinated, healthy younger adults infected with a small dose of ancestral “pre-VOC” virus (4), limiting inferences about the kinetics of emergent VOCs within the majority-vaccinated general public. Using the last known negative test in our participants, through to their first positive test, and then through serial tests up to 30 days afterwards, we were able to fully characterise viral kinetics over the course of infection rather than focusing exclusively on the initial post-onset period (7, 8). Furthermore, by linking to dates of symptom onset and conditioning on each participants estimated exposure time, we jointly—alongside each individuals viral kinetics—estimated an incubation period for each covariate. Finally, we included participants with Delta, Omicron BA.1 and BA.2 infections, enabling direct comparison between VOCs that have circulated since May 2021 in the UK (28).

Our analysis made it possible to evaluate several putative relationships between host characteristics, disease profile, infecting variant and viral kinetics. First, it has previously been suggested that both vaccination and milder or asymptomatic infection may be associated with lower viral loads and faster viral clearance (8, 30). Our finding of higher Ct values after 5+ viral exposures (whether vaccine or infection) is consistent with enhanced host immune responses due to previous exposure to spike protein. Among asymptomatic infections, there was some – albeit weak – evidence pointing to lower and earlier peak viral load. We did not find evidence that people with asymptomatic infections cleared the virus more quickly, with predicted Ct remaining below 30 for 14 days after exposure. These findings are consistent with those in the human challenge model, where viral loads remained high regardless of symptom severity (4). Within our cohort, effects of vaccine doses or symptom status on the value or timing of the peak Ct value were small, suggesting that the impact of isolation periods or likelihood of transmission would not differ based on presence of symptoms or number of previous exposures. However, we did not have data from participants with no prior exposures, limiting our ability to fully explore the role of heterogeneity in immunity.

Second, while a recent report on a large community-based study reported that Omicron BA.1 caused shorter symptomatic illness compared to the Delta variant (30), our estimated viral load kinetics did not indicate faster clearing of Omicron BA.1. This is consistent with our previously-reported data on symptom severity and duration, which found these to be the same across all VOC infections considered here (24). It has been suggested that different VOCs have altered tropism for higher or lower portions of the respiratory tract (16, 34), with BA.1 preferring upper portions and Delta those deeper. These differences in anatomical preference may alter Ct dynamics in the nasopharynx, and hence the symptom constellation may also be different with coryzal symptoms favoured in BA.1 and BA.2 infections compared to Delta (27, 35). We found a clear difference in time to limit of detection for Delta infections compared to Omicron BA.1 infections (in 35––49yo with 4 previous exposures), but the majority of people had recently received their third vaccine dose at the time of infection, and were more likely to be asymptomatic, suggesting a role of faster viral clearance for the nasopharynx in the immediate post-vaccine period (16, 22). We also found evidence of an earlier peak Ct and higher peak viral load in BA.2 infections, consistent with a recent study of serial viral loads by (9, 36) and the observed rapid spread of BA.2 across the globe during the latter stages of the BA.1 epidemic (37).

Third, the arrival of each new VOC in the UK typically resulted in a wave of infections concentrated in specific age groups, while at the same time vaccine doses have been prioritised towards older age groups (35–37). We found the peak Ct was lower in younger adults across all VOC, with older adults peaking later, and having a shorter trajectory to the LOD, despite similar vaccination doses at the time of infection. Indeed, the older adults, who received vaccine doses significantly earlier than the younger cohort, had less viral shedding. While not formally included in the model, older adults typically have an attenuated neutralising antibody response after COVID-19 vaccination (22, 23, 38). One might expect in this scenario that viral replication would therefore be more active and prolonged. Further study to understand why we observed the opposite is needed.

There are some additional limitations in our analysis. Error in Ct values is known to decrease at higher viral loads and increase at lower viral loads (5) (**Figure S2**), but to ensure model identifiability, we assumed a constant measurement error across the range of reported Ct values. Such an approximation would be particularly important to revisit if we were analysing timescales beyond the relatively short post-infection period considered here, and hence a larger range of Ct values. We also used prior exposures as a proxy to capture the potential effects of existing immunity. With more detailed linked serological and PCR data, it would be possible to examine this relationship in more detail, although this would require a substantially more complex model structure.

Previous studies inferring viral kinetics have concentrated mostly on differences between VOCs circulating at the time in question. As such, they did not consider the host of other possible confounding variables, making their results difficult to attribute solely to differences in infecting VOC (7, 13, 22, 40). In interpreting effect sizes between viral kinetic estimates, we controlled for the complex emerging picture around individual life-course exposures to SARS-CoV-2, both as infections and vaccines. Differences in numbers of infections, the VOC causing the infection, the numbers of vaccinations, and many other individual-level variables introduce many sources of possible confounding. Combining the underlying viral kinetics model with a Bayesian hierarchical framework allows this information to be jointly incorporated into the analysis. Lastly, the flexibility provided by such a framework provides a methodological foundation for future work using similar datasets. Given the ongoing antigenic turnover of SARS-CoV-2 variants of concern since late 2020, our results show it will be increasingly important to adjust for a range of changing host factors when quantifying viral kinetics and considering implications for potential infectiousness in the future.

## Acknowledgements

The authors would like to thank all the study participants, the staff of the NIHR Clinical Research Facility at UCLH including Miguel Alvarez and Marivic Ricamara. We would like to thank Bobbi Clayton, Gita Mistry, Simon Caiden and the staff of the Scientific Technology Platforms (STPs) and COVID-19 testing pipeline at the Francis Crick Institute. We thank Prof. Wendy Barclay of Imperial College and the wider Genotype to Phenotype consortium for the Alpha and Delta strains used in this study, and Max Whiteley and Thushan I de Silva at The University of Sheffield and Sheffield Teaching Hospitals NHS Foundation Trust for providing source material. We thank Prof. Gavin Screaton of the University of Oxford for the Omicron BA.1 strain used in this study.

