## Supplementary Material for "Within-host SARS-CoV-2 viral kinetics informed by complex life course exposures reveals different intrinsic properties of Omicron and Delta variants"

### Detailed description of the data

#### Cycle threshold data: ORF1ab, N and S-gene targets

For most swab tests performed during the study, multiple SARS-CoV-2 genes were targeted. ORF1ab, N-gene and S-genes were tested for. Figure S1 shows the frequency of testing for all individuals along with which genes were targeted for which individuals for each test. In order to reliably fit to as much data as available (i.e., when available, Ct values for all three targets), we investigated the relationship between the Ct values taken at the same time point as each other for the three different targets.

As shown in Figure S2, almost linear relationships across the range of Ct values for combinations of two of the three targets were observed. As such, we included a Ct target adjustment component to the statistical component of the model, whereby a linear relationship was assumed between each of the three targets and parameters of a linear regression were estimated within the same framework as the rest of the model. As such, the posteriors for the Ct target adjustment model were propagated through the rest of the inference framework. Details of the in-model adjustment used to adjust for gene target and swab type are included in the detailed methods section.

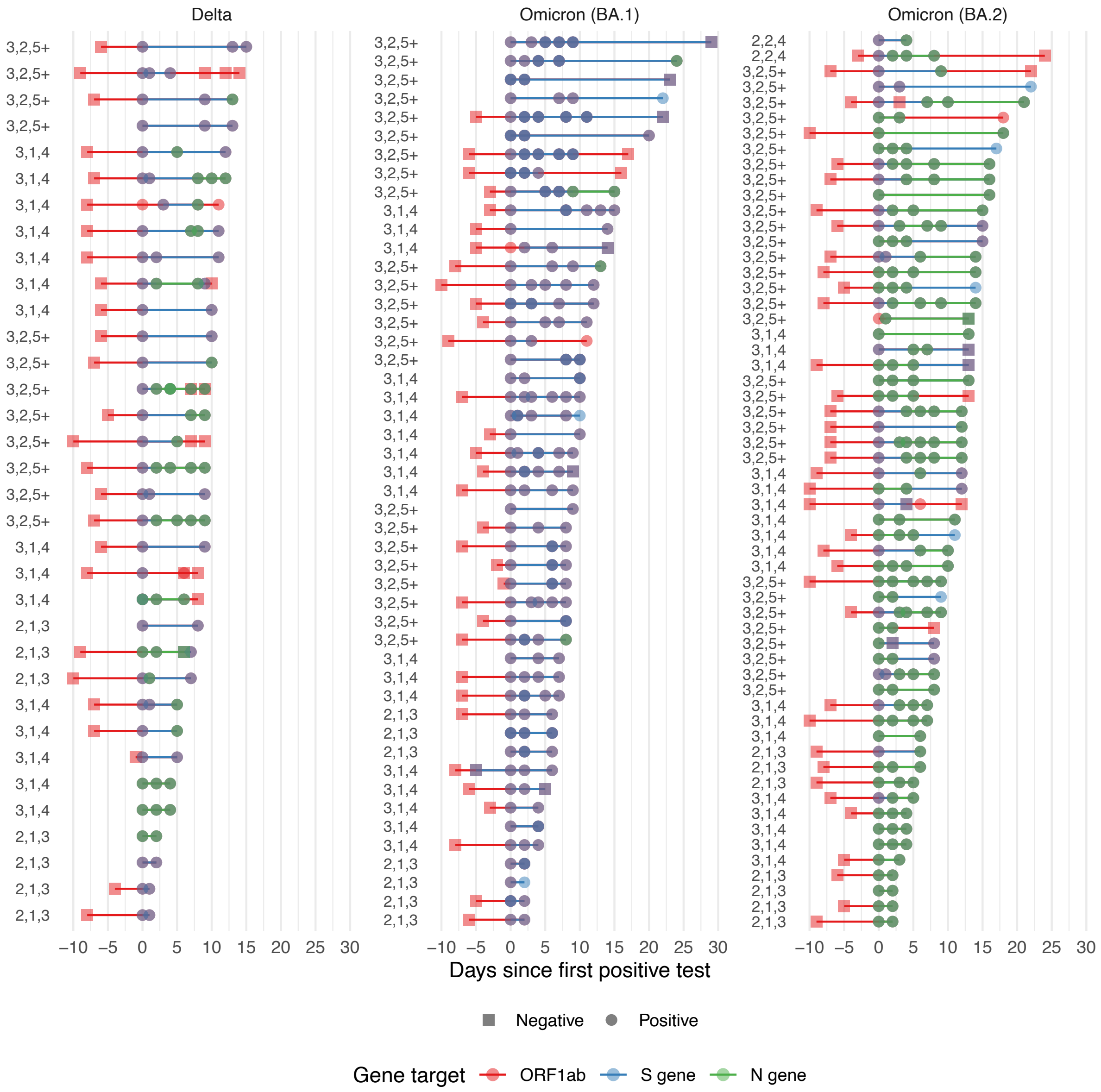

**Figure S1. Frequency of testing and which gene targets were detected for each infection episode included in the inference.** We plot the PCR test results over time to illustrate the testing frequency and which of the three potential gene targets were detected, for each infection episode with two or more positive tests. Mixed colours indicate more than one gene target was detected for a particular PCR test. The figure is of the same format as Figure 2, panel B. The only change being the colour indicates which gene targets were detected, rather than the Ct value recorded.

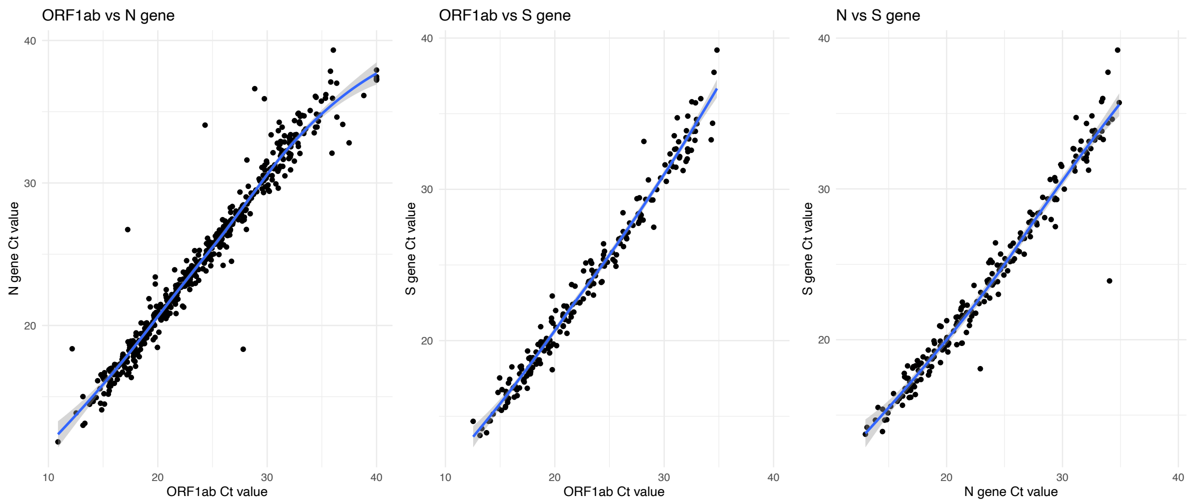

**Figure S2. Scatterplots between all combinations of the three gene targets for which Ct values were available, with fitted LOESS curves and uncertainty.** We plot the Ct values at each pair of target sites against each other to investigate their relationships and the certainty of the relationships. We do so to inform the model specification required to adjust for the gene target used.

### Detailed methods

#### Model overview and description

The biological components of the model are the *within-host viral kinetics model*, fit to longitudinal Ct data for each individual; and the *incubation period model*, fit to individual-level symptom onset data. The statistical components are a Bayesian linear regression structure used to estimate the multiplicative effect between each group of individuals and the chosen baseline set of covariates; and the hierarchical structure of the model, allowing estimation of the covariate-level effects by pooling the individual-level estimates together along with a multivariate normal distribution allowing correlations to be estimated and controlled between the individual-level parameters of the same type. Here, we give a brief technical description of each component:

- The mechanistic Ct kinetics model calculates the expected Ct at any time, given a set of individual-level parameters, including an unobserved exposure time parameter used in other terms of the likelihood function.
- The measurement model relates each observed Ct value to an expected Ct value with measurement error which we assume is normally distributed. This term is conditioned on the individual-level longitudinal Ct values.
- The component which estimates the incubation period assumes a log-normally distributed incubation period, inline with most of the previously estimates incubation periods for SARS-CoV-2. We fit shape and scale parameters for each set of covariates. This component of the model also depends on the latent exposure time parameter from the mechanistic Ct kinetics model component. This term is conditioned on the individual-level symptom onset time (relative to first positive test).
- The multiple linear regression model, which is used to regress the mechanistic model and incubation period parameters against an arbitrary selection of covariates. Specifically, effect size parameters are estimated describing the multiplicative change in the five trajectory parameters and the two incubation period parameters between covariates.
- The gene target and swab type adjustment model. We assume a linear relationship, based on plotting the raw data (Figure S2) for the adjustment model between different gene targets.
- Lastly, the hierarchical structure of the model, which see the individual-level parameters drawn from population-level distributions. The population-level

distributions are given semi-informative priors from previous studies (see Figure 1, panels B and C for a schematic of the hierarchical structure of the model). Correlations between individual-level parameters are controlled using a multivariate normal distribution.

### Viral kinetic Ct model

We used a reparameterised and reformulated version of a previously developed semi-parametric model, used in several other studies of SARS-CoV-2 viral kinetics (1–3). The model assumes a linear increase, a spline point representing the peak of viral shedding and a linear decrease on the cycle threshold scale. Given the relationship between Ct values and viral load, this represents an exponential increase and decrease (with two separate rates) on the viral load scale.

The Ct kinetics model is given by

$$g(t) = \begin{cases} c_e, & t \leq t_e \\ \dfrac{(t - t_e)(c_p - c_e)}{t_p} + c_e, & t_e < t \leq t_e + t_p \\ \dfrac{(t - t_e - t_p)(c_{\text{lod}} - c_p)}{t_{\text{lod}}} + c_p, & t_e + t_p < t \leq t_e + t_p + t_{\text{lod}} \\ c_e, & t > t_e + t_p + t_{\text{lod}}. \end{cases}$$

where $t_e$ is time the individual was exposed, $t_p$ is the time at which the individual's Ct trajectory peaks, $t_{lod}$ is the time at which the trajectory hits the limit of detection, $c_e$ is the theoretical Ct value at exposure, $c_p$ is the Ct value at the peak of the trajectory, and $c_{\text{lod}}$ is the theoretical value at which a near-perfect PCR machine would no longer detect any virus.

### Likelihood function terms

#### Viral kinetics measurement model

The log-likelihood for this component is given by

$$L_{\text{Ct}} = \log(\text{f}(x_t \mid g(t), \sigma^2) - \Phi^{-1}(0 \mid g(t), \sigma^2) + \Phi^{-1}\left(c_{\text{lod}} \mid \overline{g(t)}, \sigma^2\right)$$

where x represents the observed each Ct value is assumed to be independent and identically distributed and the variance, $\sigma^2$, is estimated in our inference framework, representing the measurement error involved in the PCR process.

The observed Ct values were censored above at Ct = 40 and truncated below at Ct = 0. The first term in the log-likelihood corresponds to expected Ct values within the observable range. The second and third terms correspond to complementary cumulative distribution functions which deal with these censored values in a relatively standard way within Bayesian inference frameworks. Specifically, $\Phi^{-1}(\mathrm{x} \mid g(t), \sigma^2)$ represents the complementary cumulative density function of the normal distribution for a given Ct value $\mathrm{x}$, with the underlying viral kinetics model at time $\mathrm{t}$ as its mean, and the same error term as the non-censored term of the likelihood function $\sigma^2$ as its variance. In the third term, corresponding to the upper censoring limit, $\overline{g(t)}$ represents expected Ct values generated by the underlying viral kinetics model that exceed the upper censoring bound. The limit of detection parameter, $c_{\mathrm{lod}}$, is a latent parameter to be estimated. This approach deals with the lower truncation and upper censoring in a statistically robust way and is an extension to previous studies which used similar underlying models but without fully accounting for censoring and truncation or modelling the underlying latent time of infection and latent limit of detection.

#### Incubation period likelihood term

To estimate the incubation period, we first of all calculate the difference between the date of the estimated infection/exposure time and the symptom onset date for each individual. We then form a censored interval of onset times  of 24 hours, given that a single date could represent any time within a 24-hour period. We denote the lower and upper limits of the censored time interval with $O_{lower}$ and $O_{upper}$, respectively.

We assume a log-normally distributed incubation period, a common parametric choice when estimating the incubation period of SARS-CoV-2 (4). We then use the following term contributing to the overall log-likelihood function for estimating the parameters, $\mu_{IP}$ and $\sigma_{IP}$, of the log-normal distribution:

$$L_{IP} = \log(F_{IP}(O_{\text{upper}} \mid \mu_{IP}, \sigma^2_{IP}) - F_{IP}(O_{lower} \mid \mu_{IP}, \sigma^2_{IP})),$$

Where the $F_{IP}$ terms denote the cumulative distribution function of the log-normal distribution, $\mu_{IP}$ and $\sigma_{IP}$ correspond to the location and scale parameters of the log-normal distribution, which we estimate in our framework, and the $O_{\text{upper}}$ and $O_{\text{lower}}$ terms refer to the lower and upper bounds on the censored interval of the time of symptom onset, which given the high resolution of our dataset, is 1 day for each individual.

### Covariate-level priors

#### Viral kinetics model

The following distributions were chosen as priors for the covariate-level parameters of the viral kinetics model (i.e. the population-level parameters in the hierarchical structure)

$$
\begin{aligned}
&t_e \sim \text{Normal}(t_{\text{bound}} + 5,5) \\
&t_p \sim \text{Normal}(\log(5), 0.5) \\
&t_{\text{lod}} \sim \text{Normal}(\log(10) + t_p, 0.5) \\
&c_e \sim \text{Normal}(50, 5) \\
&c_p \sim \text{Normal}(\text{logit}^{-1}(0), 1) \\
&c_{\text{lod}} = c_e
\end{aligned}
$$

where an individual-level value $t_{\text{bound}}$ represents the censored interval in which infection must have occurred, based on each individual's symptom onset time (relative to their first positive test). Specifically, $t_{\text{bound}} = \max(-t_{\text{onset}}, 0)$.

These weakly-informative priors were chosen by combining estimates from the human challenge study (Killingley et al. (2022) (5)) and estimates of the viral kinetics from previous studies (Hay et al. (2022) (1)). We have little prior knowledge of each individual's exposure time, which is why the prior for this parameter at the population level was non-informative. The other parameters have relatively strong evidence from previous studies, which is how we justify using semi-informative priors. Alternative choices for the individual-level priors and the correlation between the individual-level parameters are analysed and discussed in the *Sensitivity analysis with alternative model fits* section (**Figures S9—S12**).

#### Incubation period model

The following distributions were chosen as priors for the covariate-level incubation period parameters

$$
\begin{aligned}
&I_{\text{mu}} \sim \text{Normal}(1.621,0.0640) \\
&I_{\text{sigma}} \sim \text{Normal}(0.418, 0.0691)
\end{aligned}
$$

,

here $I_{\text{mu}}$ and $I_{\text{sigma}}$ are the mean and standard deviation of the assumed log-normal distribution of the latent incubation period. As these are parameters of a log-normal distribution, the values given should be interpreted on the exponential scale. Doing so gives the probability that symptom onset relative to the estimated time since exposure. For example, the mean of the prior for $I_{\text{mu}}$ corresponds to an incubation period of approximately 5 days since exposure, when exponentiated and interpreted on the natural scale (i.e., $\exp(1.621) \approx 5.06$ days).

### Individual-level priors

#### Viral kinetics model

The individual-level parameters are given by

$$
\begin{aligned}
t_e^{ind} &= \exp(\beta_{t_e} t_e + \eta_{t_e}) \\
t_p^{ind} &= \exp\left(\beta_{t_p} t_p + \eta_{t_p}\right) \\
t_{lod}^{ind} &= \exp(\beta_{t_{lod}} t_{lod} + \eta_{t_{lod}}) \\
c_e^{ind} &= \text{logit}^{-1}(\beta_{c_e} t_e + \eta_{c_e}) \\
c_p^{ind} &= \text{logit}^{-1}(\beta_{c_p} c_p + \eta_{c_p}) \\
c_{lod}^{ind} &= c_e^{ind} \\
\beta_i &\sim \text{Normal}(0, 0.2)
\end{aligned}
$$

where the interpretation of the parameters is equivalent to the description in the covariate-level parameters section. The superscript *ind* refers to the fact these are the individual-level parameters. The timing parameters $t_e^{ind}$, $t_p^{ind}$ and $t_{lod}^{ind}$ are fit on an exponential scale, and the Ct value parameters $c_e^{ind}$ and $c_p^{ind}$ are fit on the logit scale. Given that the priors for the covariate-level parameters are given above, to fully define the individual-level parameter priors, we need only define the priors for the effect size parameters (the $\beta_i$ terms) and the individual-level variation matrix $\eta$.

#### Individual-level variation

We model residual individual variation across process model parameters ($\eta_p^{\text{ind}}$, where $p = t_e, t_p, t_{\text{lod}}, c_e, c_p$) as potentially correlated (i.e. an individual with a higher peak Ct value may also be more (or less) likely to have a higher Ct value at the limit of detection than an individual with a lower peak Ct value) using a multivariate normal distribution,

$$\eta_p^{\text{ind}} \sim \text{MVN}(0, \boldsymbol{\Sigma})$$

Where $\Sigma$ is a 5 x 5 covariance matrix which we decompose for computational stability into a diagonal matrix containing process parameter specific scale parameters ($\Delta$) and a symmetric correlation matrix ($\Omega$) as follows,

$$\boldsymbol{\Sigma} = \boldsymbol{\Delta}\,\boldsymbol{\Omega}\,\boldsymbol{\Delta}$$

$$\boldsymbol{\Delta} = \begin{pmatrix} \sigma_{t_e} & 0 & 0 & 0 & 0 \\ 0 & \sigma_{t_p} & 0 & 0 & 0 \\ 0 & 0 & \sigma_{t_{lod}} & 0 & 0 \\ 0 & 0 & 0 & \sigma_{c_e} & 0 \\ 0 & 0 & 0 & 0 & \sigma_{c_p} \end{pmatrix}$$

$$\boldsymbol{\Omega} = \begin{pmatrix} 1 & \omega_{t_e, t_p} & \omega_{t_e, t_{lod}} & \omega_{t_e, c_e} & \omega_{t_e, c_p} \\ \omega_{t_p, t_e} & 1 & \omega_{t_p, t_{lod}} & \omega_{t_p, c_e} & \omega_{t_p, c_p} \\ \omega_{t_{lod}, t_e} & \omega_{t_{lod}, t_p} & 1 & \omega_{t_{lod}, c_e} & \omega_{t_{lod}, c_p} \\ \omega_{c_e, t_e} & \omega_{c_e, t_p} & \omega_{c_e, t_{lod}} & 1 & \omega_{c_e, c_p} \\ \omega_{c_p, t_e} & \omega_{c_p, t_p} & \omega_{c_p, t_{lod}} & \omega_{c_p, c_e} & 1 \end{pmatrix}$$

Where $\omega_{p_1, p_2}$ dictates the correlation between process parameters. We use a Lewandowski-Kurowicka-Joe (LKJ) prior for the symmetric correlation matrix **Ω** and a weakly informative half-normal prior for the scale parameters,

$$\boldsymbol{\Omega} \sim \text{LKJCorr}(\upsilon)$$
$$\sigma_p \sim Half - Normal(0, 0.2)$$

For the LKJ prior $\upsilon = 1$ results in a uniform prior over all correlations, $\upsilon < 1$ places more weight on larger correlations and $\upsilon > 1$ places more weight on small amounts of correlations. By default, we set $\upsilon = 1$. On top of this potentially correlated process parameter model, we also model the extreme scenario where we assume no correlation between process parameters. I.e. $\omega_{p_1, p_2} = 0$ for all parameter combinations.

As the multivariate normal density and LKJ prior on correlation matrices both require their matrix parameters to be factored we have parameterised our model directly in terms of Cholesky factors of correlation matrices using the multivariate version of the non-centered parameterisation, as recommended by the Stan User Manual. This increases numerical stability and sampling efficiency, reducing the computational requirements of model fitting.

#### Bayesian linear regression structure

We added a linear regression structure on top of the existing mechanistic and statistical modelling framework, allowing us to flexibly regress against any number of covariates within our dataset, all within the same inference structure. Therefore, the inference included estimating multiplicative effect sizes, which were used in post-fitting to adjust the covariate-level estimates (Figures 3 and 4) for each covariate. This has not previously been implemented in similar models.

#### Gene target and swab type adjustment

Using the same programmatic structure as the linear regression used to regress against the covariates of interest, we adjusted for which gene targets were detected and which swab type was used (wet or dry).

#### Individual-level posterior and posterior predictive distributions

The parameters of the mechanistic part of the model were fit for each individual. Using these estimated individual-level parameters, we are able to simulate likely Ct trajectories and check the goodness-of-fit of the model against the observed Ct value data. We plot the three timing posterior distributions (**Figure S3**), the Ct peak value posterior distributions (**Figure S4**) and the posterior predictive distributions (**Figure S5**) with the underlying data we fit to: symptom onset and Ct values at the three gene targets investigated.

#### Computational details

We implemented the model using R 4.2.2 (7) and CmdStan 2.3.1 (8). We ran the model for 3000 iterations, after 1000 warm-up iterations. We used the standard R-hat < 1.05 diagnostic to assess convergence of the chains for each model parameter. The model fit took no longer than a few minutes on a 2021 M1 MacBook Pro, passed all the in-built standard diagnostic tests and fit with no divergent transitions.

### Timing parameters

### Delta-infected individuals

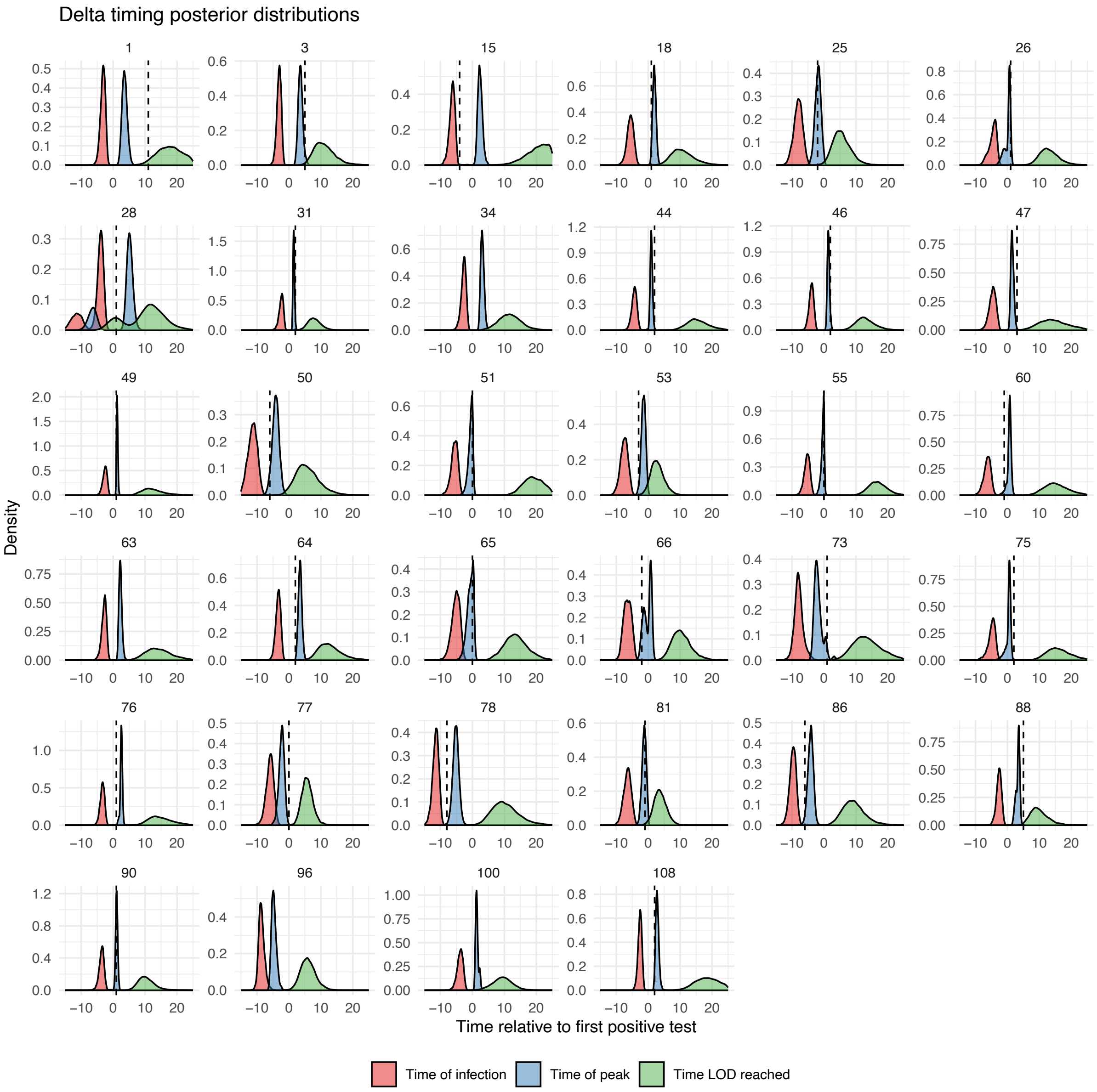

**Figure S2.** Posterior distributions for the individual-level time of infection, time at which the peak Ct value is reached and the time the LOD is reached for individuals infected with the Delta variant. The IDs match the other individual-level posterior plots (**Figures S2—S8**). All times are relative to each individual's first positive test. Dashed vertical lines represent the time at which symptoms began for each individual, where reported.

### Omicron (BA.1)-infected individuals

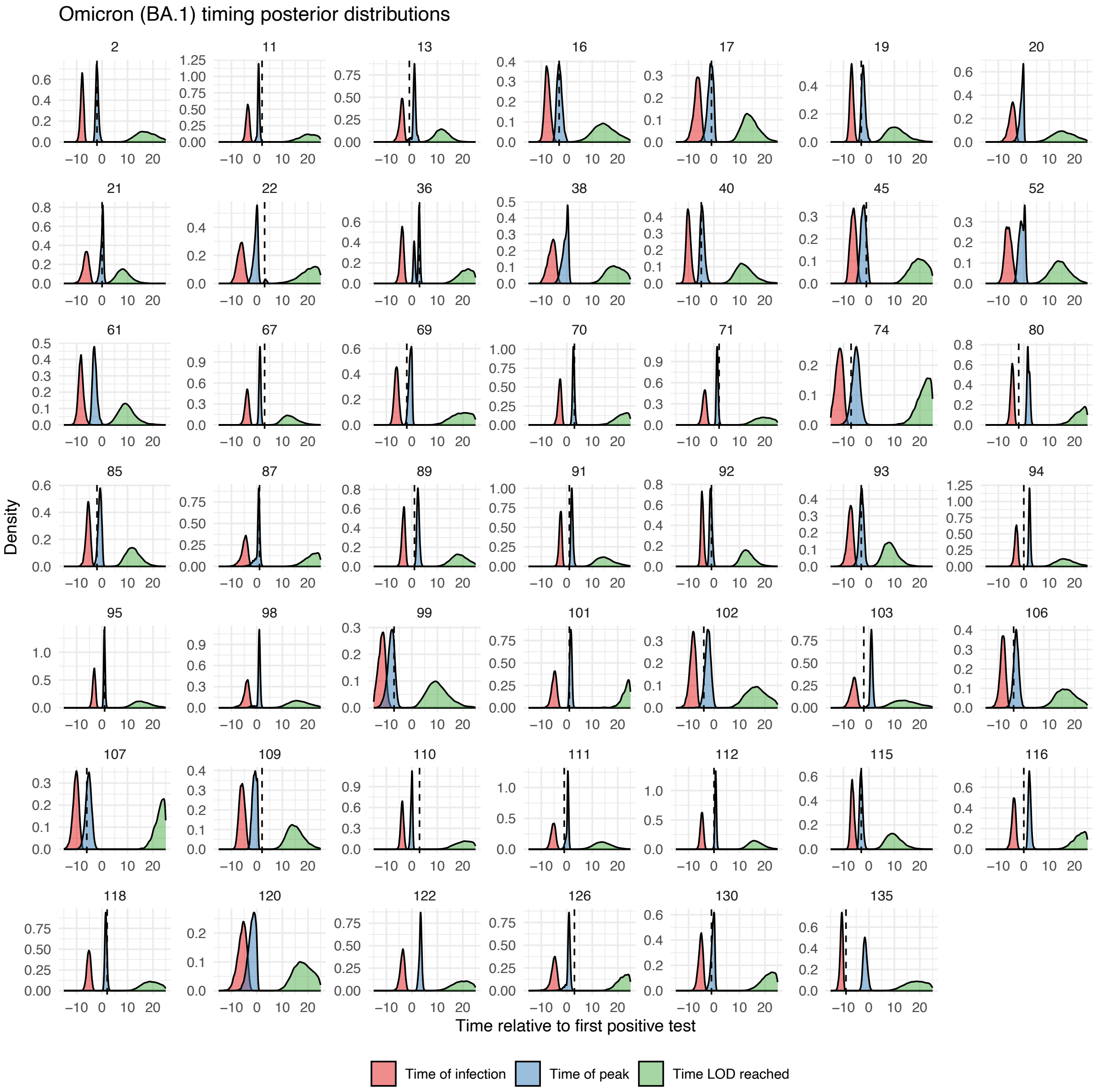

**Figure S3.** Posterior distributions for the individual-level time of infection, time at which the peak Ct value is reached and the time the LOD is reached for individuals infected with the Omicron BA.1 variant. The IDs match the other individual-level posterior plots (**Figures S2—S8**). All times are relative to each individual's first positive test. Dashed vertical lines represent the time at which symptoms began for each individual, where reported.

### Omicron (BA.2)-infected individuals

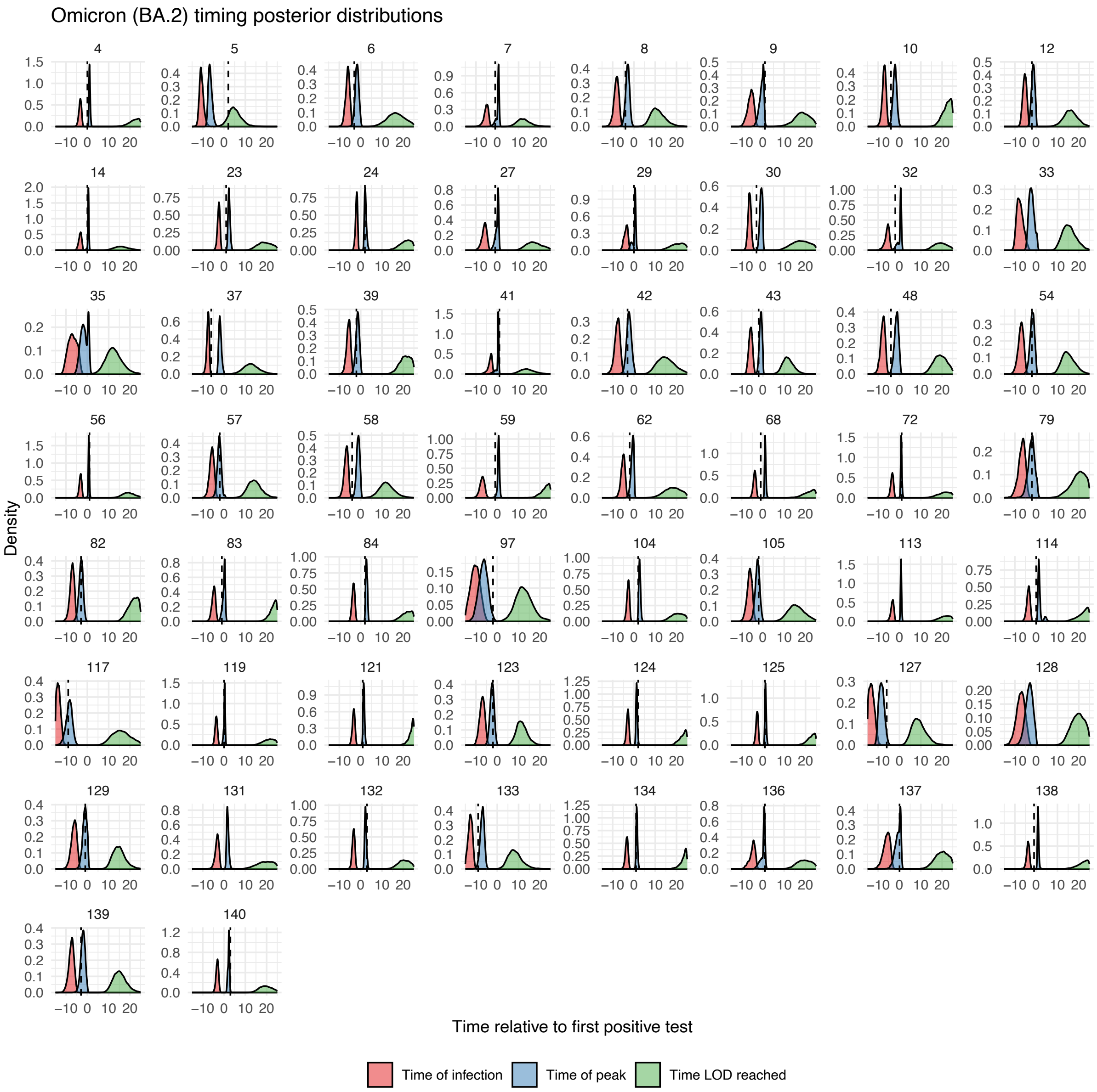

**Figure S4.** Posterior distributions for the individual-level time of infection, time at which the peak Ct value is reached and the time the LOD is reached for individuals infected with the Omicron BA.2 variant. The IDs match the other individual-level posterior plots (**Figures S2—S8**). All times are relative to each individual's first positive test. Dashed vertical lines represent the time at which symptoms began for each individual, where reported.

### Ct value parameters

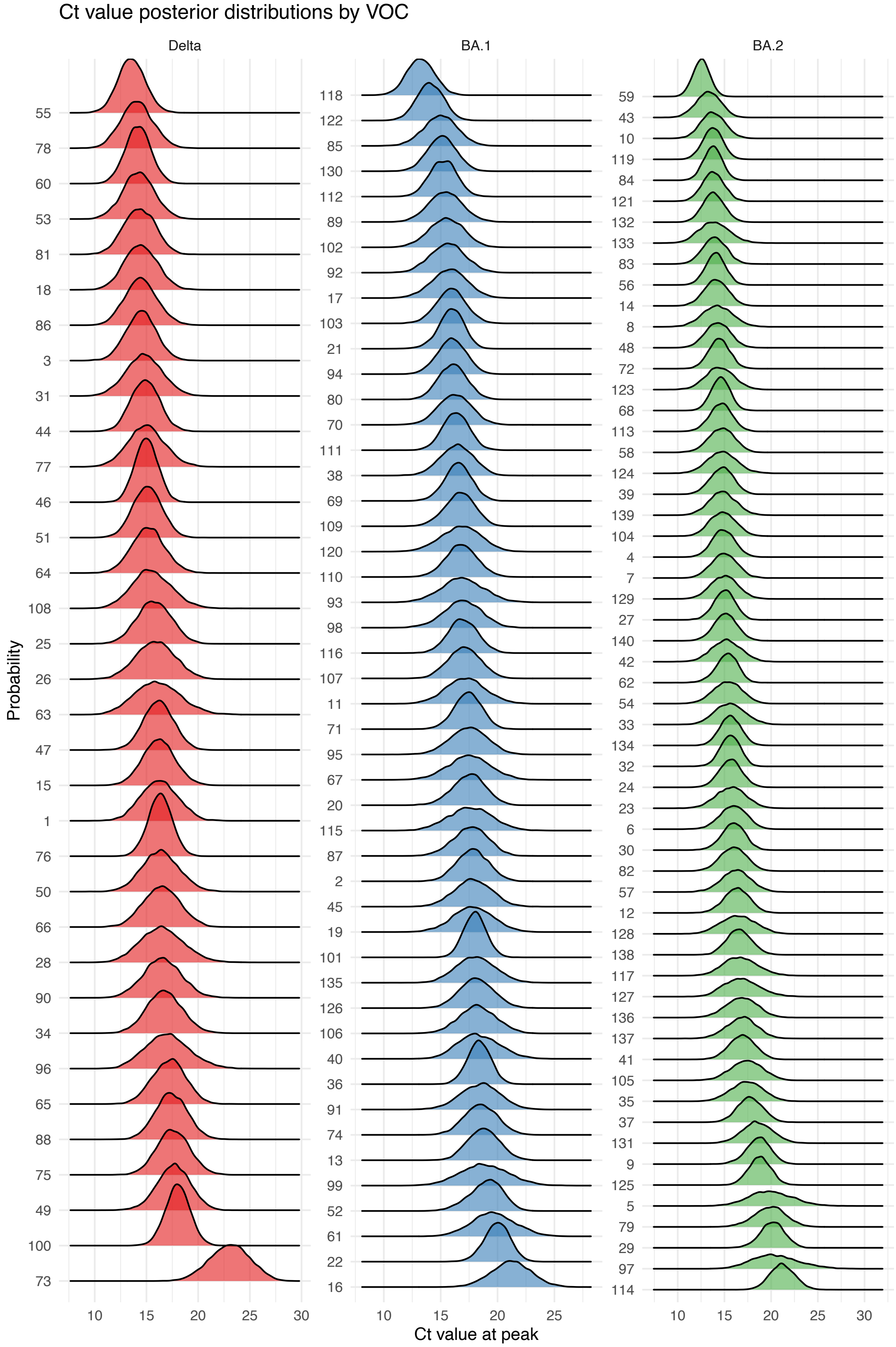

**Figure S5.** Posterior distributions for the individual-level peak Ct value, with IDs which match the other individual-level posterior plots (**Figures S2—S8**).

### Viral kinetics model fits

We present the fitted Ct trajectories stratified by the three VOCs under investigation. The fitted trajectories presented are generated using the fitted individual-level parameters for each individual.

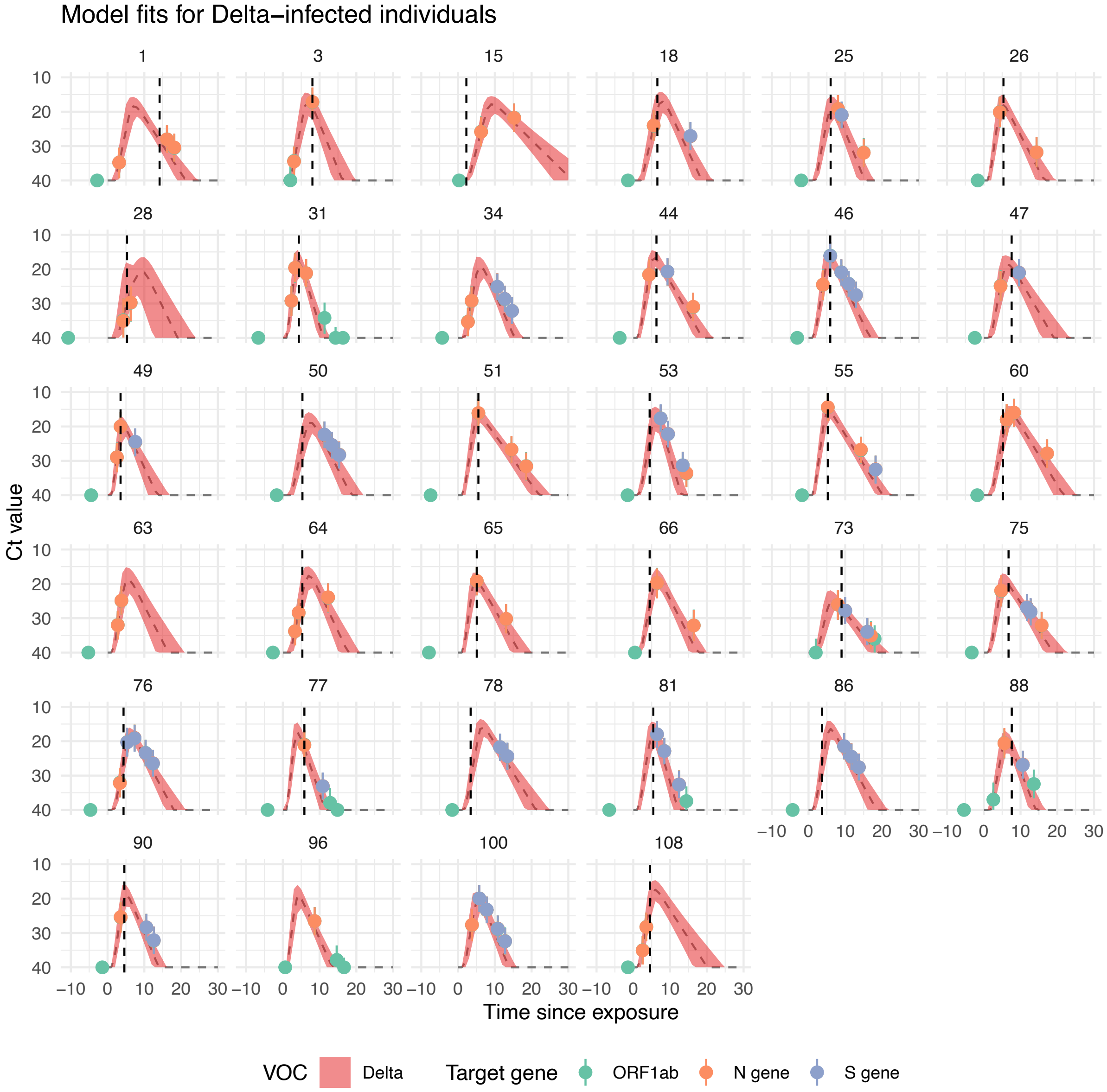

**Figure S6.** Posterior predictive distributions for all Delta-infected individuals, produced by simulating the Ct trajectory model using the inferred posterior distributions shown in **Figures S2—S5**, with matching IDs. All times are relative to each individual's estimated time of exposure. Dashed vertical lines represent the time at which symptoms began for each individual, where reported.

### Omicron (BA.1)-infected individuals

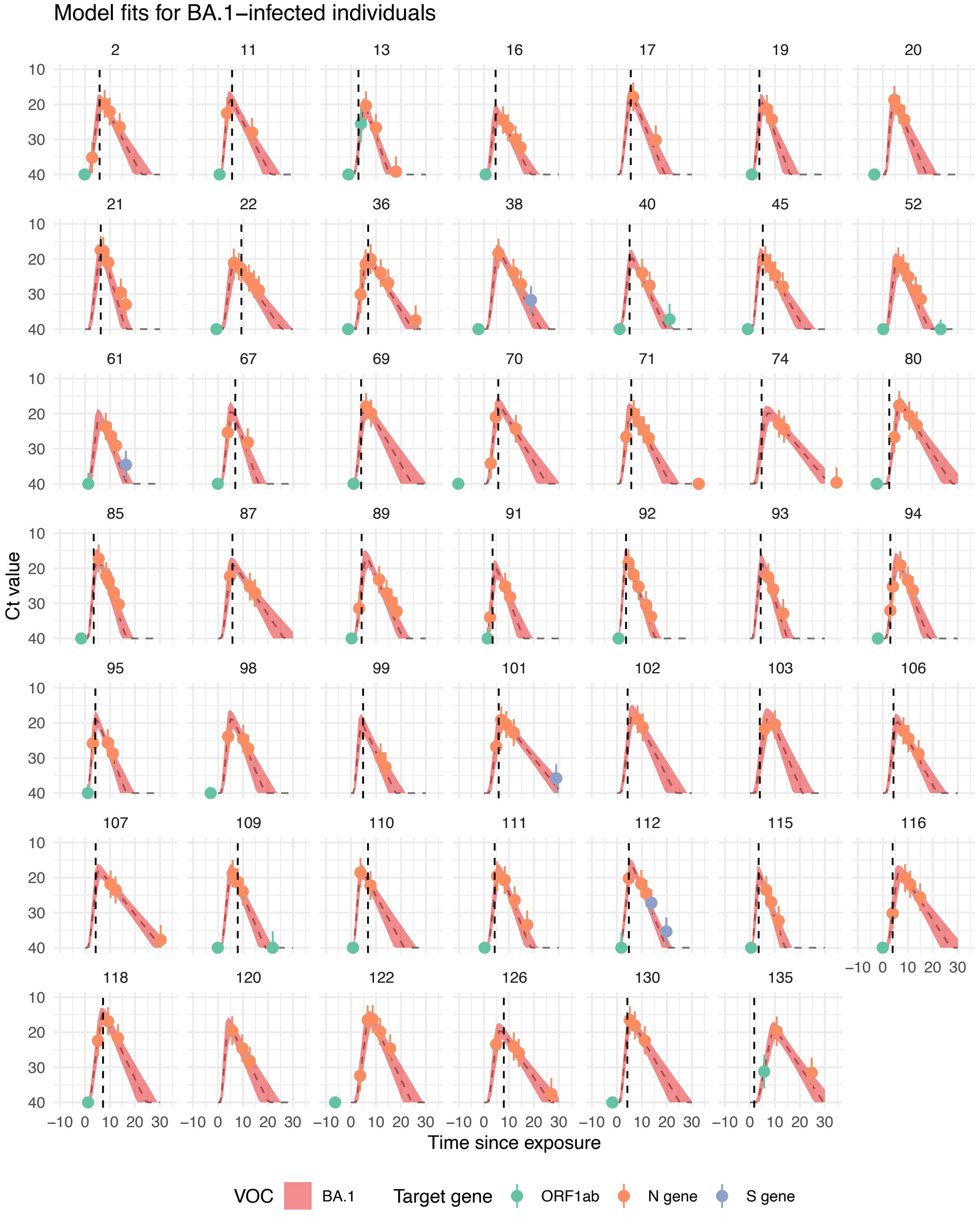

**Figure S7.** Posterior predictive distributions for all Omicron (BA.1)-infected individuals, produced by simulating the Ct trajectory model using the inferred posterior distributions shown in **Figures S2—S5**, with matching IDs. All times are relative to each individual's estimated time of exposure. Dashed vertical lines represent the time at which symptoms began for each individual, where reported.

### Omicron (BA.2)-infected individuals

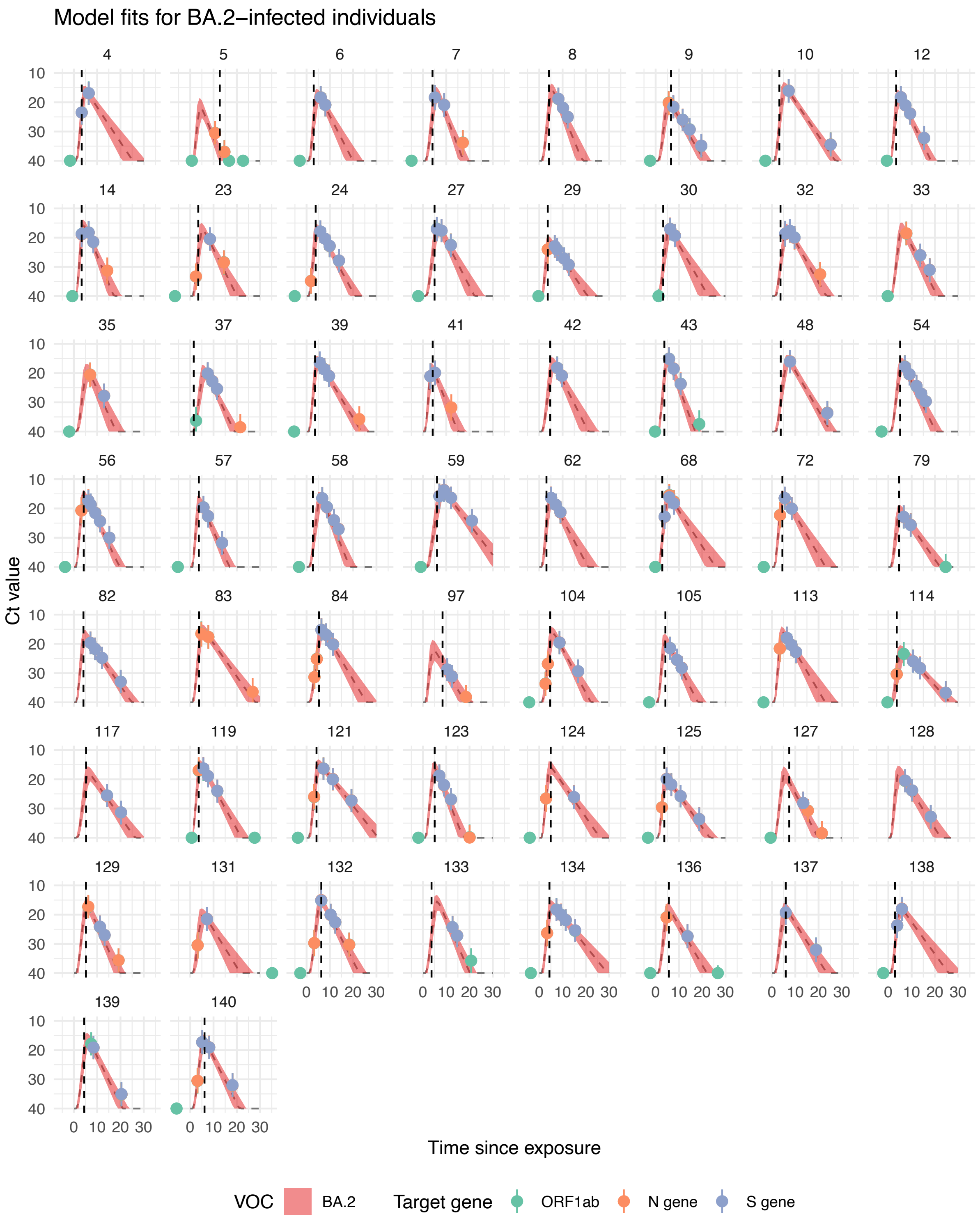

**Figure S8.** Posterior predictive distributions for all Omicron (BA.2)-infected individuals, produced by simulating the Ct trajectory model using the inferred posterior distributions shown in **Figures S2—S5**, with matching IDs. All times are relative to each individual's estimated time of exposure. Dashed vertical lines represent the time at which symptoms began for each individual, where reported.

### Timing of rapid test positivity

For rapid test positivity (assumed Ct = 30), baseline trajectories crossed the positivity threshold at 3.4 (95% CrI: 2.4—4.2) days after exposure and would have remained positive until 15.4 (95% CrI: 13.2—17.1) days after exposure. We estimated that Delta trajectories would turn rapid test positive at an almost identical time to the baseline trajectories (for symptomatic BA.1 infections in 34-49yo with 4 previous exposures) but would only remain positive until 14.1 (95% CrI: 11.8—15.8) days after exposure. We estimated that BA.2 trajectories would turn positive 3.1 (95% CrI: 2.1—3.9) days after exposure and would remain positive until 16.1 (95% CrI: 13.8—17.9) days after exposure. Trajectories for all other covariates would turn positive at almost identical times to the baseline trajectories. However, we estimated substantial variation in the duration of rapid test positivity for the other covariate categories. Trajectories would become rapid test negative for participants with 3 or 5+ post exposures sooner than baseline individuals; specifically at 13.6 (95% CrI: 11.1—15.7) and 12.3 (95% CrI: 10.4—13.9) respectively. Lastly, we found that trajectories for participants in the 20–34 and 50+ age groups would become negative later than for reference individuals, at 16.0 (95% CrI: 13.4—18.3) and 16.7 (95% CrI: 13.8—19.1) days after exposure, respectively.

### Tables of population-level posterior estimates

| | VOC | | |
|---|---|---|---|
| | **Delta** | **BA.1 (baseline)** | **BA.2** |
| **Peak Ct value** | 15.1 (13.8—16.5) | 16.1 (14.9—17.2) | 15.0 (13.9—16.1) |
| **Timing of the peak (days)** | 6.0 (4.9—7.3) | 5.6 (4.7—6.5) | 5.2 (4.4—6.2) |
| **Timing of the LOD (days)** | 19.6 (15.5—24.9) | 24.9 (20.5—30.8) | 26.4 (21.9—32.1) |

**Table S1. Table of the median and 95% credible intervals of the population-level Ct model parameters by VOC.**

| | Symptom status | |
|---|---|---|
| | **Symptomatic (baseline)** | **Asymptomatic** |
| **Peak Ct value** | 16.1 (14.9—17.2) | 17.0 (15.3—18.7) |
| **Timing of the peak (days)** | 5.6 (4.7—6.5) | 5.2 (4.0—6.7) |
| **Timing of the LOD (days)** | 24.9 (20.5—30.8) | 22.8 (17.6—29.5) |

**Table S2. Table of the median and 95% credible intervals of the population-level Ct model parameters by symptom status.**

| | Total number of exposures | | |
|---|---|---|---|
| | **3** | **4 (baseline)** | **5+** |
| **Peak Ct value** | 16.7 (15.1—18.3) | 16.1 (14.9—17.2) | 17.9 (16.5—19.4) |
| **Timing of the peak (days)** | 5.5 (4.3—6.9) | 5.6 (4.7—6.5) | 5.2 (4.2—6.6) |
| **Timing of the LOD (days)** | 20.8 (16.2—26.9) | 24.9 (20.5—30.8) | 19.6 (15.0—25.1) |

**Table S3. Table of the median and 95% credible intervals of the population-level Ct model parameters by total number of antigenic exposures.**

| | Age | | |
|---|---|---|---|
| | **20—34** | **35—49 (baseline)** | **50+** |
| **Peak Ct value** | 16.3 (15.1—17.6) | 16.1 (14.9—17.2) | 17.7 (16.4—19.0) |
| **Timing of the peak (days)** | 4.2 (3.5—5.1) | 5.6 (4.7—6.5) | 5.2 (4.0—6.6) |
| **Timing of the LOD (days)** | 30.2 (25.0—37.1) | 24.9 (20.5—30.8) | 31.2 (24.4—40.9) |

**Table S4. Table of the median and 95% credible intervals of the population-level Ct model parameters by age.**

### Sensitivity analysis with alternative model fits

To check the sensitivity of the statistical and mechanistic components of our model to different covariates and prior distributions, we ran a number of model variations. Specifically, we re-fit our model considering four different changes to model structure in turn: omission of all covariates other than VOC; increasing the potential for variation to be attributed to individual-level effects; removing the estimation of correlation structure between individual-level responses and removing symptom onset data from the likelihood.

### No other covariates other than VOC

When the model was fitted with only VOC as a covariate, we observed similar trends in effect size across VOCs, but with substantially different and more varied point estimates for Ct value at peak and narrower credible intervals. This reflects our finding that additional covariates are likely to influence viral dynamics – such as prior exposures that would have generated immunity – and omission of these covariates can result in too much of the individual-level variation being attributed to the VOC rather than shifting population characteristics over time.

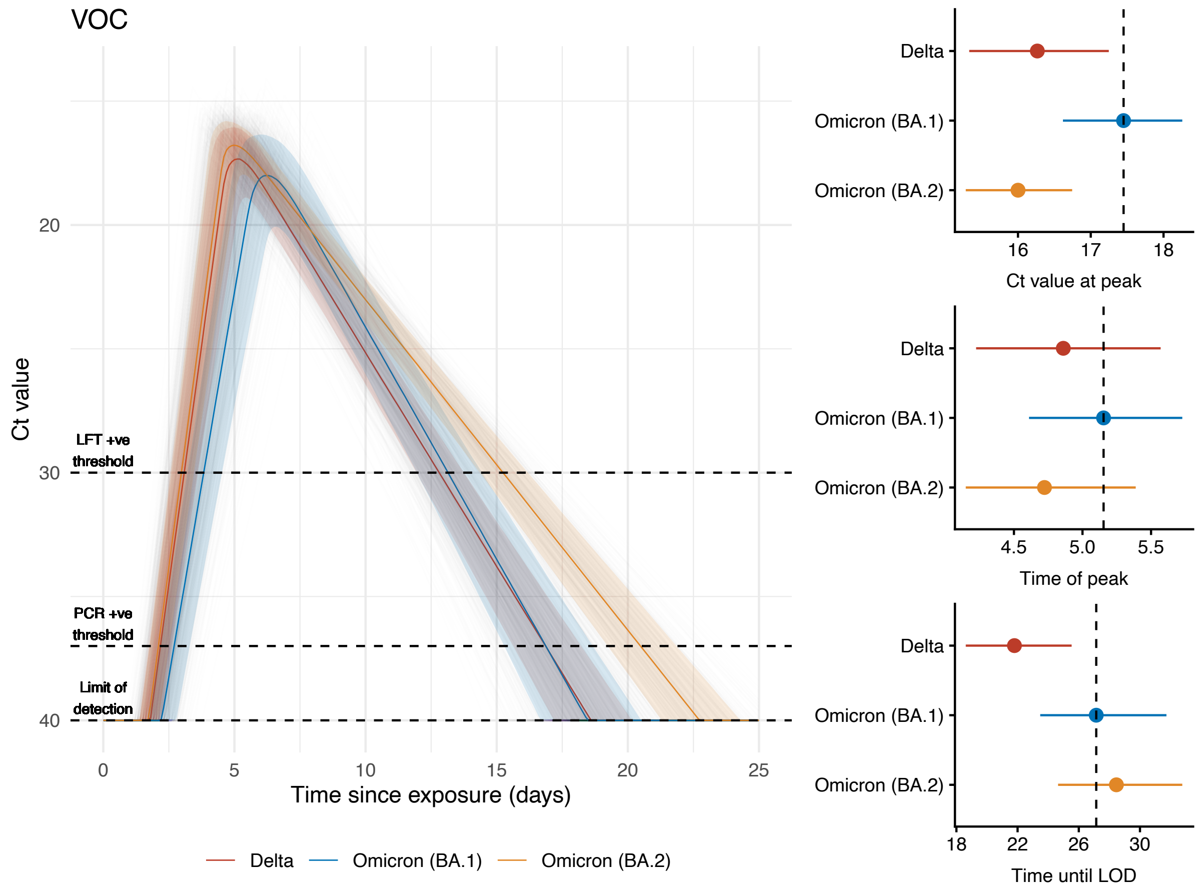

**Figure S9.** Population-level fit for the model when only the infecting VOC was used as a covariate.

### Greater individual-level variation

When we changed the prior distribution assumptions to allow for greater individual-level variation in response, our estimates had larger uncertainty, but similar findings with regards to the effect of prior exposures and age, indicating that these population-level characteristics still had a discernible effect against a background of noisy individual-level responses.

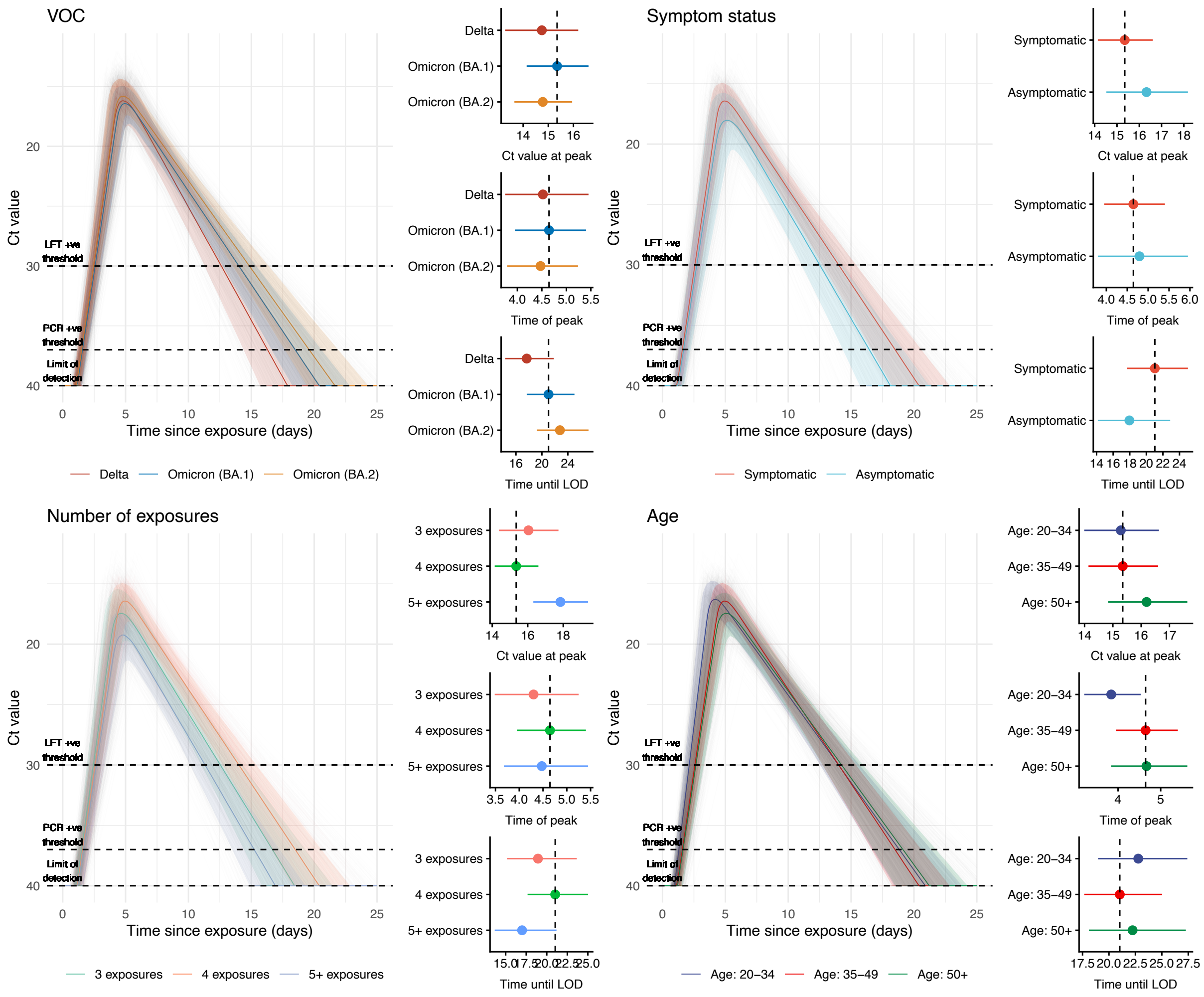

**Figure S10.** Population-level fits for the model when substantially higher levels of individual-level variation were permitted.

### No correlation between individual-level parameters

To assess how much individual-level parameters were jointly informed by the assumed correlation structure, we fitted the model assuming no correlation between individual-level parameters. We obtained similar estimates to our main analysis, again suggesting that population-level characteristics had a discernible effect even when individual-level dynamics were fitted with a prior representing no correlation.

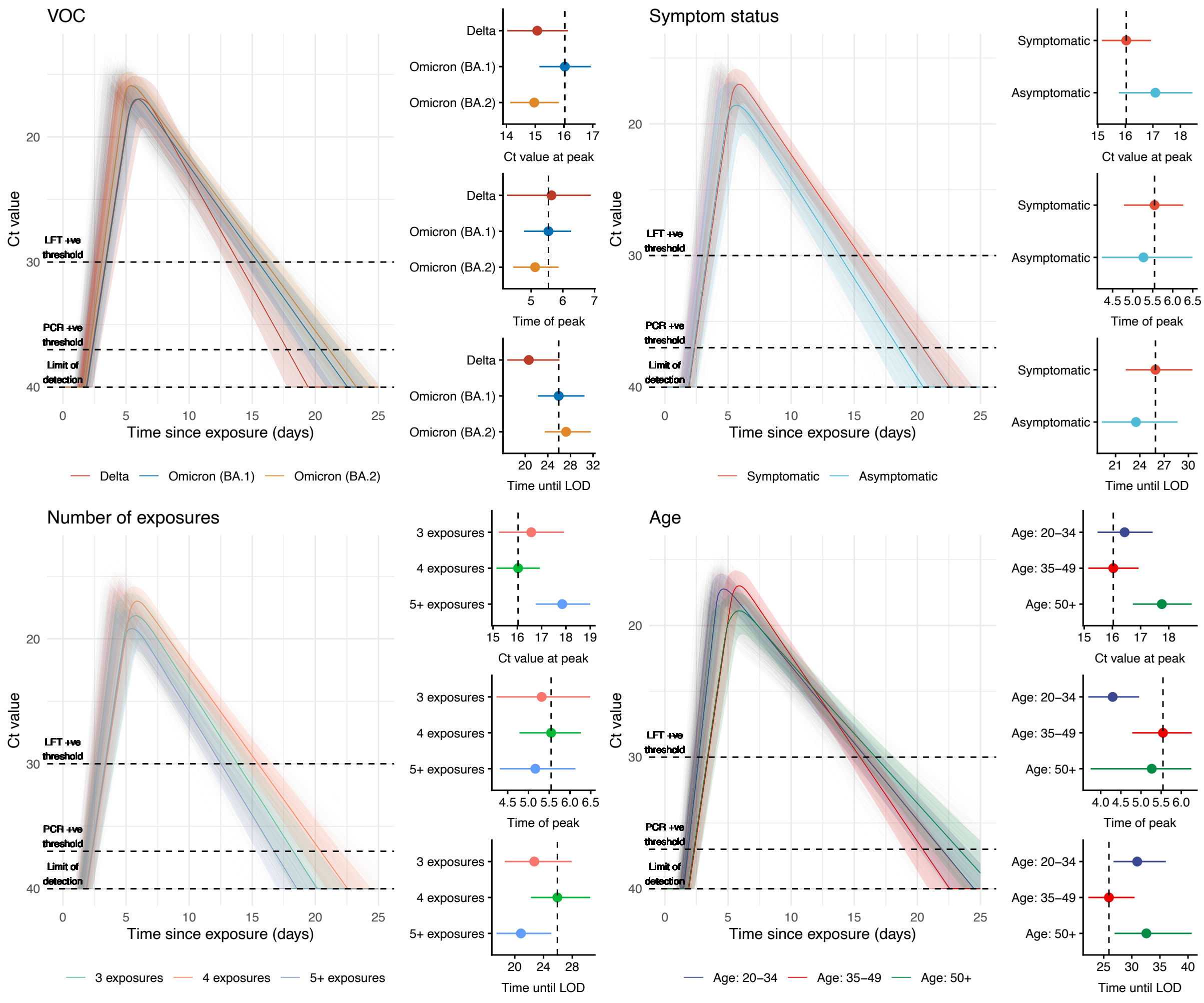

**Figure S11.** Population-level fits for the model when parameterised with a prior that almost entirely removes correlation between the individual-level parameters, controlled by the multivariate normal distribution structure. Specifically, the parameter of the LKJ-prior used to control the multivariate normal distribution was set to a far higher value ($\eta = 50$), whereby high values correspond to lower and lower levels of permitted correlation between individual-level parameters.

### No symptom onset data used in likelihood

When we omitted symptom onset data from the likelihood, we obtained similar conclusions about the role of prior exposures and age, but lower point estimates for the time until peak Ct value. This suggests that symptom onset data – anchored to infection time via our prior on the incubation period – is important for supplementing the sparsity in early post-infection Ct measurements and its interaction with fast-changing Ct values early in the infection.

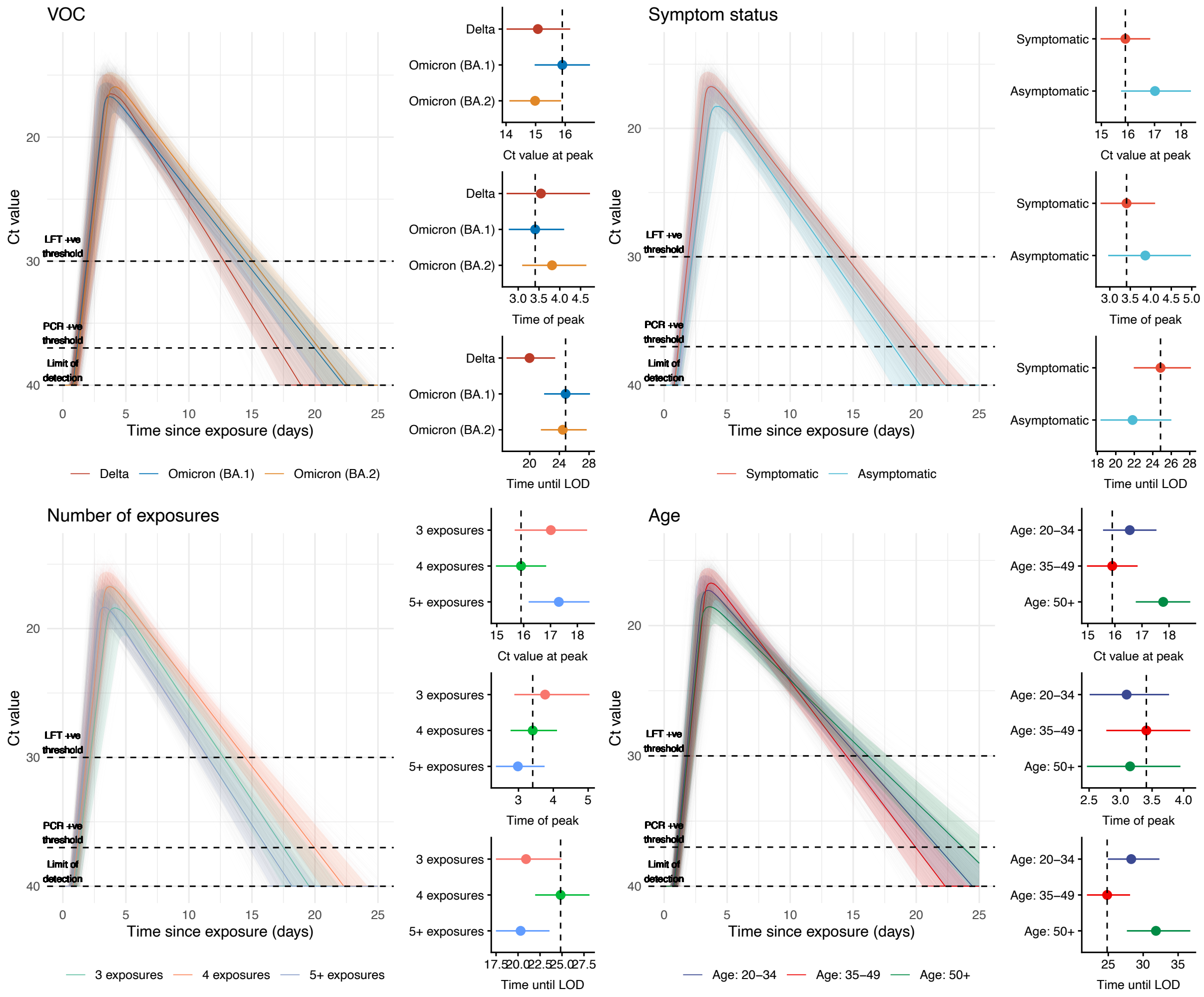

**Figure S12.** Population-level fits for the model when symptom onset data was omitted from the fitting. The time of exposure for each individual is informed in the model presented in the main text by a combination of the Ct trajectory model, fit to Ct value data, and the incubation period model, fit to symptom onset data. We refit the model after removing the symptom onset component of the likelihood, arriving at the above population-level fits.

### Alternative Ct threshold for Figure 4

To investigate the effect of the assumed Ct value threshold used in Figure 4 (Ct = 20), we varied the Ct threshold (Ct = 25) and replotted Figure 4.

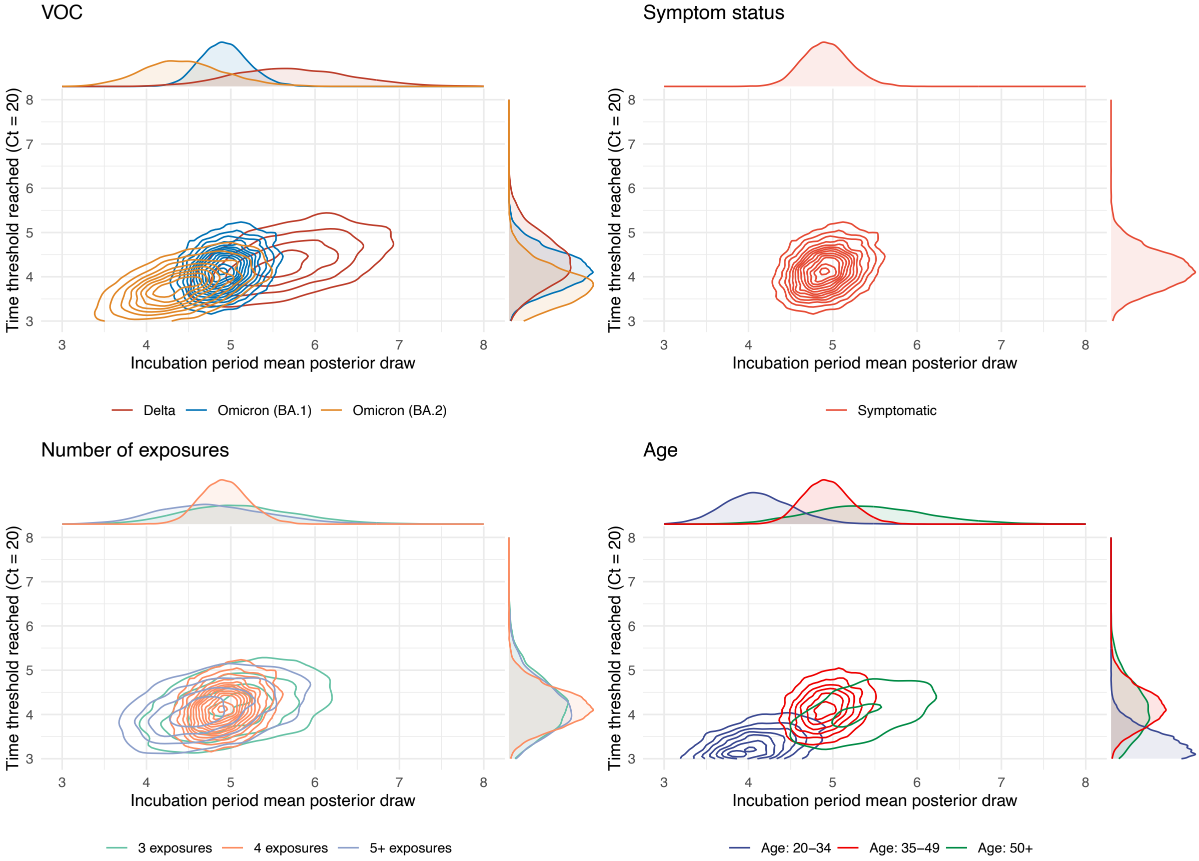

**Figure S13. Bivariate density plots of median incubation periods against the time at which trajectories surpassed an assumed Ct value (Ct = 25), for each covariate. All panels**: We replot **Figure 4** in the main text with a different assumed Ct value threshold (Ct = 25 vs Ct = 20). **A.** Parameter values for the three VOCs considered. **B.** Parameter values corresponding to symptomatic infections. **C.** Parameter values corresponding to the numbers of exposures considered. **D.** Parameter values for the age groups considered.

### Incubation period estimates

To investigate differences in the overall incubation period estimates, sampled using both the inferred mean and standard deviation parameters, are given, stratified by each covariate included in the main analysis.

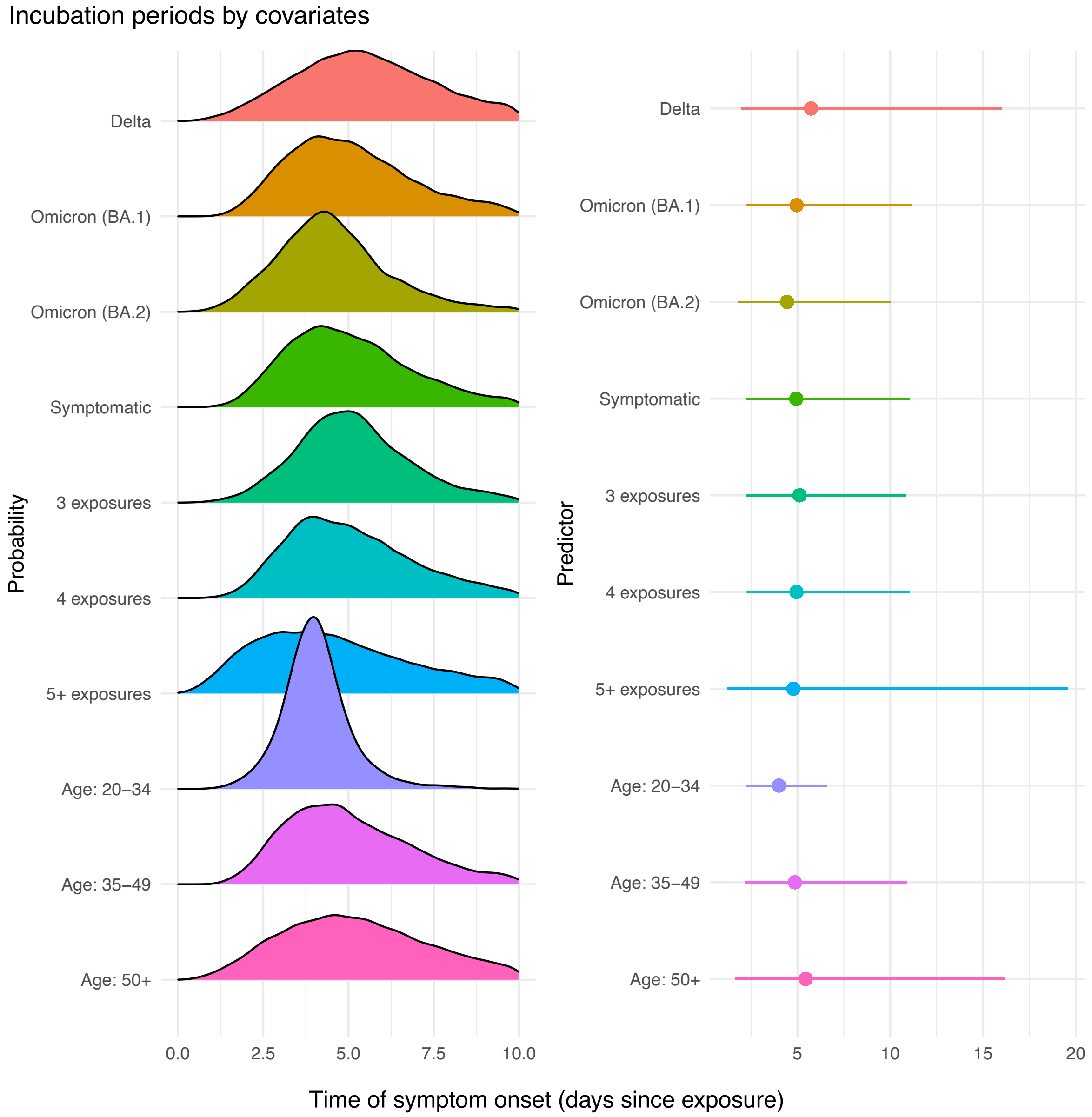

**Figure S14. Density and effect size plots of the inferred incubation periods, for each covariate. A**: Incubation periods generated using the inferred posterior distributions for both the mean and standard deviation, stratified by each covariate. **B.** The mean (dots) and 95% CrI of the incubation periods plotted in **Panel A.**

### Gene target and swab type adjustment posteriors

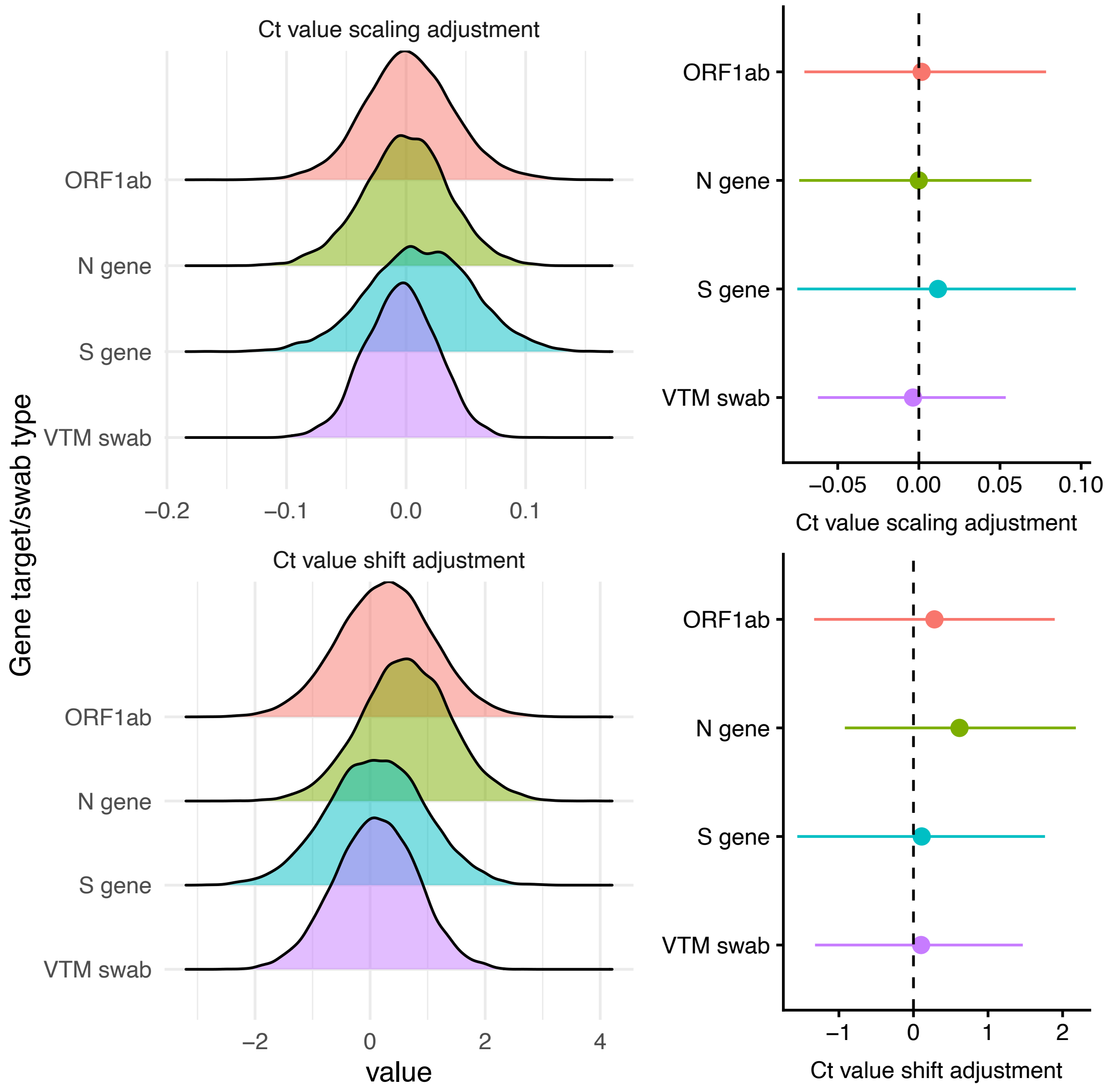

**Figure S15. Posterior density plots for the estimated effect sizes for the Ct adjustment component of the model.** We plot the posterior distributions (left panel) and the median and 95% credible interval (right panel) for the adjustment factors for the Ct scale and Ct shift parameters, stratified by each Ct target and swab type.

### Population-level prior vs posterior comparison

To directly investigate the influence of our choice of priors and our model (conditioned on our dataset) used in the main results, we plot the population-level priors (equivalent for each predictor) against the inferred posteriors, by the predictors used in the main results. The inferred posterior distributions clearly differ from the priors in location and width for all parameters and covariates, showing that the posterior distributions are strongly informed by the data.

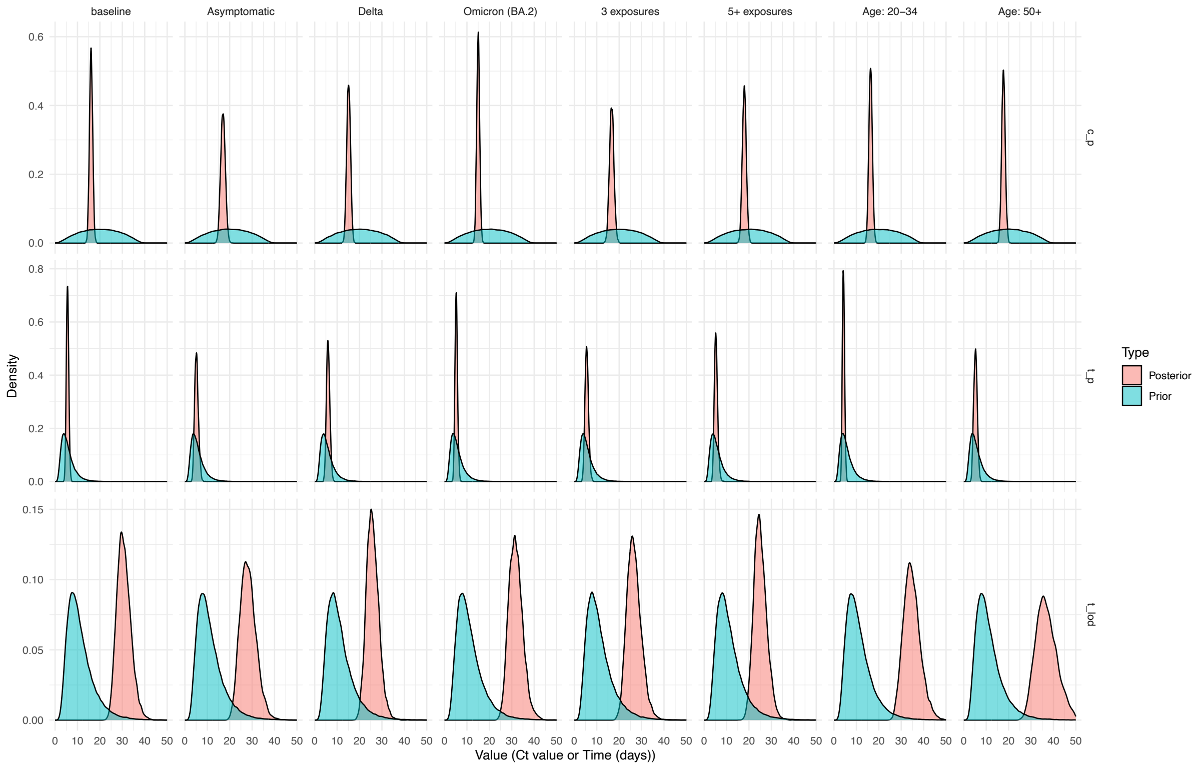

**Figure S16. Population-level priors and inferred population-level posterior distributions for peak Ct value ($c_p$), time to peak Ct value ($t_p$), and time to LOD ($t_{lod}$) by each covariate used in the main results.**

8. Documentation [Internet]. stan-dev.github.io. [cited 2023 Mar 24]. Available from: //mc-stan.org/users/documentation/
